# Monitoring sodium content in packaged foods sold in the Americas and compliance with the Updated Regional Sodium Reduction Targets

**DOI:** 10.1101/2024.05.23.24307787

**Authors:** Yahan Yang, Nadia Flexner, Maria Victoria Tiscornia, Leila Guarnieri, Adriana Blanco-Metzler, Hilda Núñez-Rivas, Marlene Roselló-Araya, Paola Arévalo-Rodríguez, Maria Fernanda Kroker-Lobos, Francisco Diez-Canseco, Mayra Meza-Hernández, Kiomi Yabiku-Soto, Lorena Saavedra-Garcia, Lorena Allemandi, Leo Nederveen, Mary R. L’Abbé

## Abstract

**Background:** Sodium reduction is a cost-effective measure to prevent noncommunicable diseases. The World Health Organization (WHO) established a target of a 30% relative reduction in mean population intake of sodium by 2025. The Pan American Health Organization (PAHO) published sodium reduction targets for packaged foods in 2015, expanding and updating the targets in 2021 to help Member States with its efforts in reducing population sodium intake.

**Objective:** This study examined the current sodium levels in packaged foods among five countries in the Americas and monitored cross-sectional and longitudinal compliance with the sodium targets from 2015 to 2022.

**Methods:** Food labels were systematically collected from the main supermarkets in five countries in the Americas region in 2022. Sodium levels per 100g and per kcal for collected food labels in 16 PAHO categories and 75 subcategories were analyzed and compared against the Updated Regional Targets. Further analysis of three countries that have longitudinal data for 2015-2016, 2017-2018 and 2022 was conducted to compare sodium per 100 g against the 2015 PAHO Targets.

**Results:** A total of 25,569 food items were analyzed. Overall, *‘processed meat and poultry’* had the highest sodium levels, although there were large variations within categories. 47% and 45% of products met the sodium per 100g and per kcal 2022 targets, respectively. Peru had the highest compliance, whereas Panama had the lowest for both targets. Among Argentina, Costa Rica and Peru, the proportion of foods meeting the 2015 PAHO lower targets were 48, 53 and 61% for 2015-2016, 2017-2018 and 2022, respectively.

**Conclusions:** This study showed that around half of the examined foods met their respective sodium targets and there have been small improvements in compliance over time. Further efforts are required to reach the WHO’s global sodium reduction goal by 2025, such as implementation of mandatory sodium reduction targets and front-of-pack labelling regulations.

## 1. Introduction

Hypertension is one of the major risk factors for cardiovascular diseases (CVDs) and it has been estimated to account for 10.8 million deaths globally in 2019^1,2^. The number of adults with hypertension has been increasing in the past few decades, with more than 1 billion adults affected by hypertension in 2019, globally^3^. It has been well demonstrated that excessive consumption of sodium is a significant causal risk factor for the development of hypertension and reducing dietary sodium can have a favorable effect on the cardiovascular system^4^. The World Health Organization (WHO) published a guideline in 2012 recommending less than 5g of salt (2g of sodium) intake per day for adults^5^. However, the global average salt intake in 2019 was estimated to be more than double the recommendation at 10.8g of salt (4.3g of sodium) per day^1^.

In 2013, WHO established an action plan that included reducing salt intake by 30% by 2025 and all 194 Member States signed up^6^. A series of interventions had been proposed, including reformulation of foods to contain less sodium, establishing public food procurement policies to limit foods high in sodium, using front-of-package labelling that inform consumers of high sodium products and using mass media campaigns to reduce sodium intake^7^. However, a report released in early 2023 showed that the world is off-track to achieve this global target, with only 5% of countries (Brazil, Chile, Czech Republic, Lithuania, Malaysia, Mexico, Saudi Arabia, Spain and Uruguay) implementing at least two mandatory and comprehensive sodium reduction policies and all WHO sodium-related ‘best buys’^1^.

To support the Americas region in accomplishing the global target, the Pan American Health Organization (PAHO) published a set of regional sodium reduction targets (2015 PAHO targets) in 2015 for 18 food categories that were commonly sold in the region^8^. In 2019, a study examined the sodium levels in packaged foods sold in 14 Latin American and Caribbean countries (LAC) according to the 2015 PAHO targets. The study found 82% compliance with the regional target and 47% compliance with the lower and stricter targets^9^. The regional target level was the maximum level (mg/100g) or upper limit, whereas food manufacturers were encouraged to reach the lower target level that is reflective of the average level in reference countries^8^. A later study published in 2021 showed that compliance with regional targets increased from 83% to 89% from 2015-2016 to 2017-2018 among four Latin American countries ^10^. Therefore, PAHO updated the targets in 2021 to set stronger regional sodium reduction targets for 2022 and 2025 (2022 and 2025 PAHO targets) which are similar to the 2015 PAHO lower targets, and it further categorized foods into 16 main categories and 75 subcategories^11,12^. Similarly, WHO published global benchmarks, defined as the lowest maximum value for each subcategory from existing national or regional targets, for sodium levels in foods across different food categories^13^. However, there has been no further follow-up study monitoring sodium levels in packaged foods in the Americas since 2018. Thus, the objectives of this study were to 1) examine the current sodium levels among five countries in the Americas (Argentina, Canada, Costa Rica, Panama, and Peru), as well as monitor compliance with the Updated Regional Sodium Reduction Targets for 2022; and 2) monitor the progress of sodium reduction among three Latin American countries that have longitudinal data.

## 2. Methods

This cross-sectional study was conducted using data from five countries: Argentina, Canada, Costa Rica, Panama, and Peru. Data from the Nutrition Facts table (NFt) (n=44,570) were collected in supermarkets in Argentina (n=4,340), Costa Rica (7,402), Panama (n=1,509) and Peru (n=5,378) between March and August 2022. The Food Label Information Program for Latin America and Caribbean countries (FLIP-LAC) was used for data collection and analyses. FLIP-LAC is a smartphone-based technology and web database, and the methodology was developed by The University of Toronto (U of T)^14^. The Canadian data (n=25,941) were extracted from the Food Label Information and Price (FLIP) database collected in 2020^14^.

Foods were classified into 16 major categories and 75 subcategories described in the Updated PAHO Regional Sodium Reduction Targets^12^. The updated PAHO target categorization was individually completed by each country team and the U of T research team validated the results and resolved discrepancies with country team members. The sodium content was standardized into mg/100g and mg/kcal where data were available. Foods excluded from the analyses included duplicated items, foods that could not be categorized under the PAHO categories (n=19,001), and foods that have 0 kcal were additionally excluded from the mg/kcal analysis, due to mathematical restrictions (n=387). Summary statistics were calculated for food products by food categories and by country. The sodium level was compared against the Updated PAHO Regional Sodium Reduction Targets (**Appendix A**) to determine the proportion of foods that met or exceeded the target level set for 2022.

Since three countries in this study had baseline data from 2015-2016 and 2017-2018, categorized under the 2015 PAHO targets, a further sub-analysis was conducted by categorizing the foods collected in 2022 from Argentina, Costa Rica and Peru into the 2015 PAHO target categories (18 food categories)^8^. Sodium levels were compared against the 2015 PAHO regional (upper) target level as well as the lower target level. Proportion of foods meeting the regional targets was compared among the three timepoints. All analyses were conducted with R studio (5.12.10) and Microsoft Excel (2016).

## 3. Results

After exclusions, this study included a total of 25,569 items across 16 PAHO sodium reduction main categories and 75 subcategories (Argentina n=2,515, Canada n=15,268, Costa Rica n=3,875, Panama n=1,121, and Peru n=2,790).

### 3.1 Sodium levels per 100g and per kcal by PAHO categories

Of the 23,663 foods that had sodium per 100g available, *‘processed meat and poultry’* and *‘sauces, dips, gravy, and condiments’* had the highest median sodium levels per 100g (both at 800mg/100g), followed by *‘fats and oils’* (720mg/100g) (**Table 1**). The variations within categories were high, particularly among *‘sauces, dips, gravy and condiments’* (SD = 5,733mg/100g), *‘processed fish and seafood’* (SD = 972mg/100g) and *‘processed vegetables, beans and legumes’* (SD = 599mg/100g). There were variations between the median sodium levels between countries, with the largest variation within *‘sauces, dips, gravy, and condiments’*, *‘soy products and meat alternatives’* and *‘processed meat and poultry’*. The lowest variation between countries was within *‘fresh or dried plain pasta and noodles’*, *‘processed fish and seafood’* and *‘ready-made foods’*.

**Table 1.**
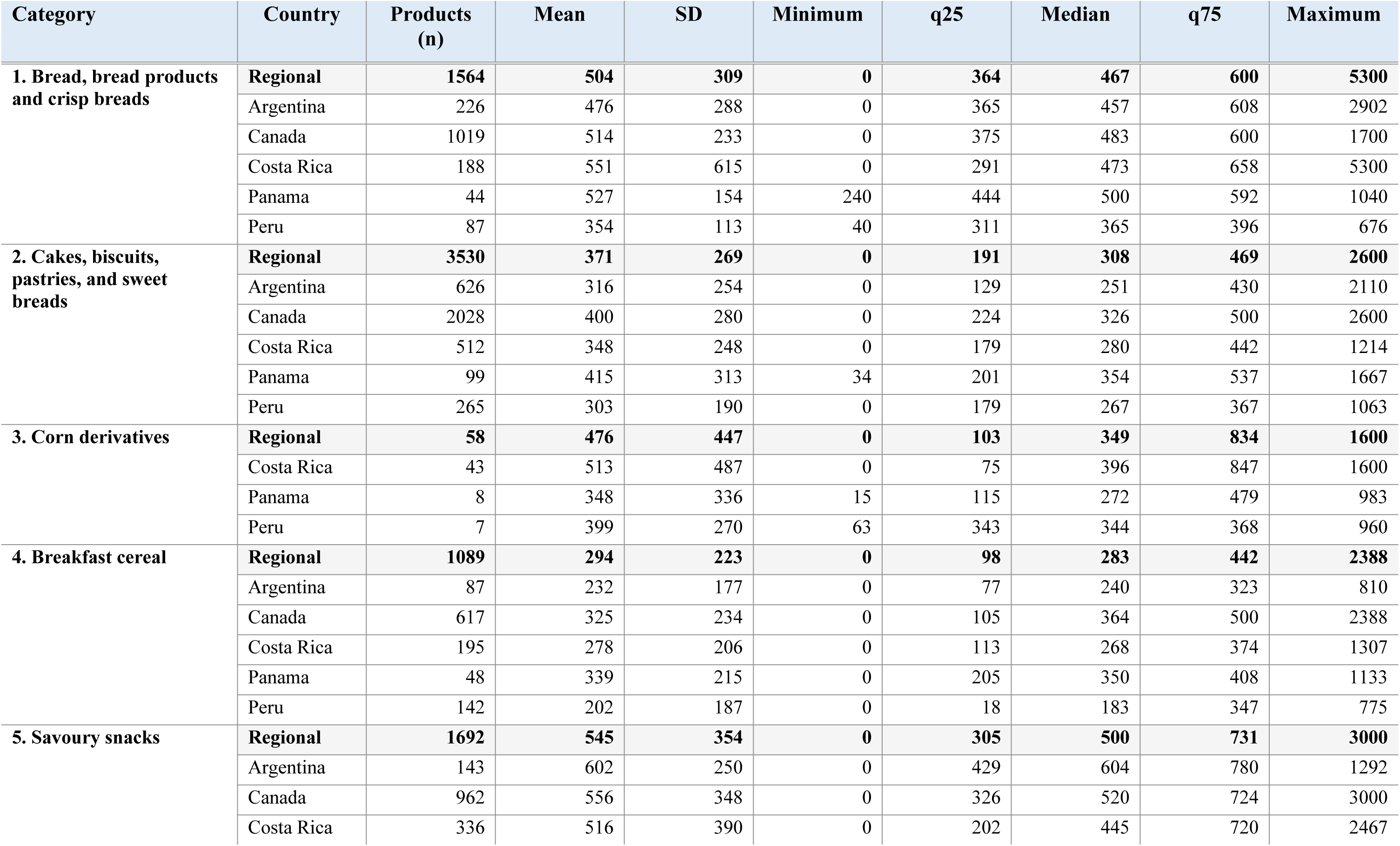
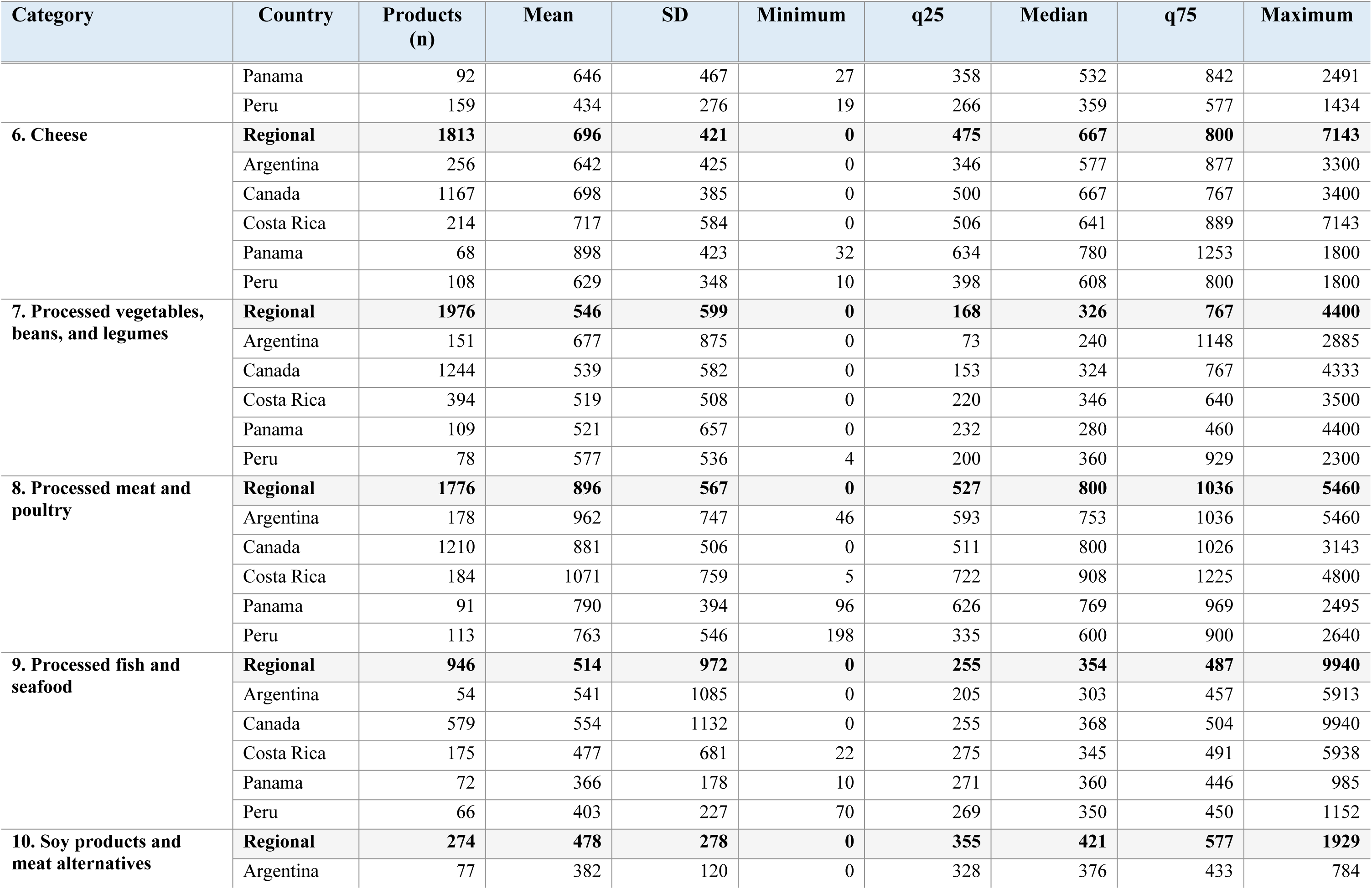
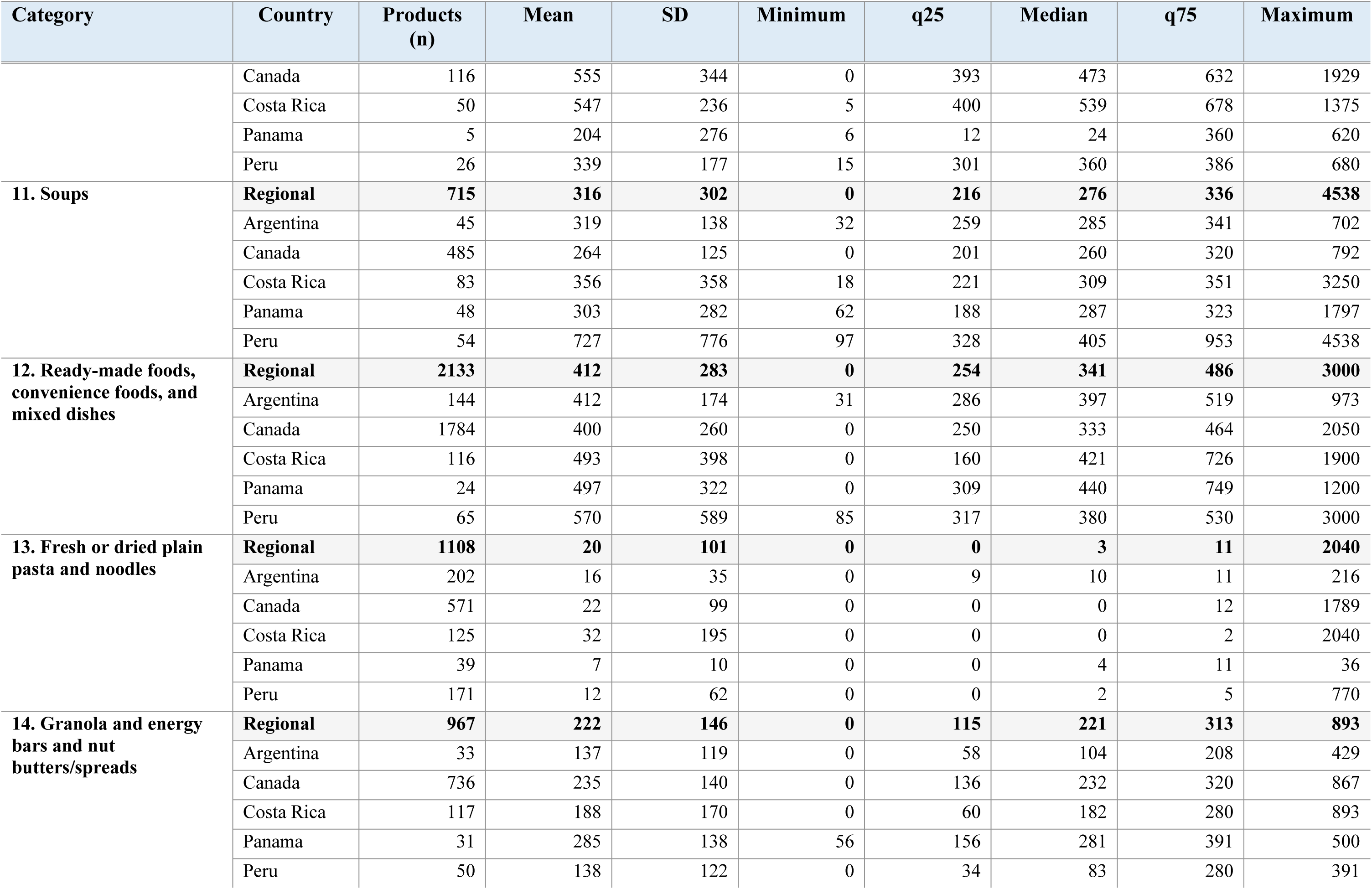
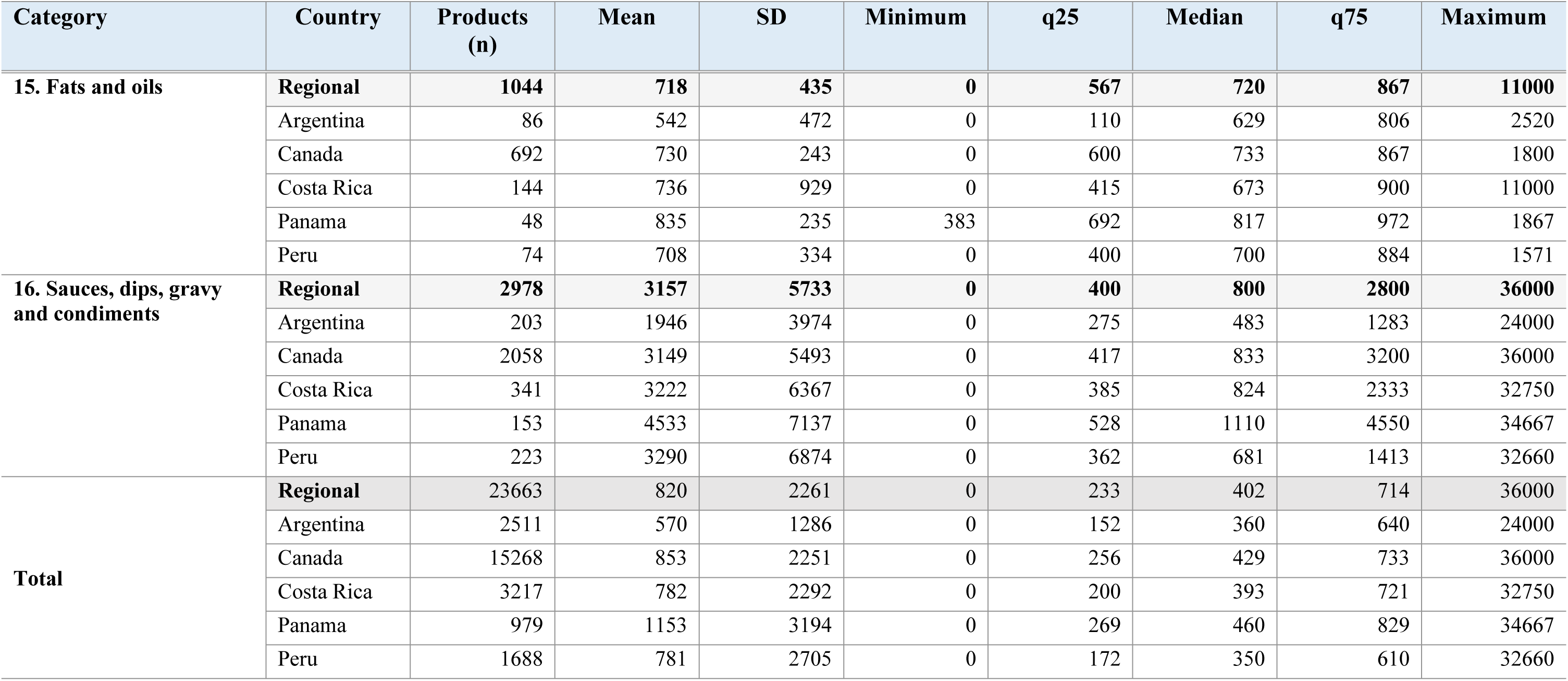
Distribution of sodium content per 100g/ml of packaged foods per PAHO food category at the regional level and by country.

Of the 23,236 foods that had sodium per kcal data available, *‘sauces, dips, gravy, and condiments’*, *‘soups’* and *‘processed vegetables, beans, and legumes’* have the highest median sodium level per kcal of 7.8mg/kcal, 6.0mg/kcal and 5.1mg/kcal, respectively (**Table 2**). The lowest median sodium levels per kcal were among *‘fresh or dried plain pasta and noodles’* (0mg/kcal), *‘granola and energy bars and nut butters/spreads’* (0.5mg/kcal) and *‘cakes, biscuits, pastries, and sweet breads’* (0.7mg/kcal). The largest variations between countries were found among *‘sauces, dips gravy and condiments’*, *‘soups’* and *‘soy products and meat alternatives*. Variation within category was highest among *‘sauces, dips, gravy, and condiments’* (SD=41.4mg/kcal), *‘processed vegetables, beans and legumes’* (SD=20.2mg/kcal) and *‘soups’* (SD=17.8mg/kcal).

**Table 2.**
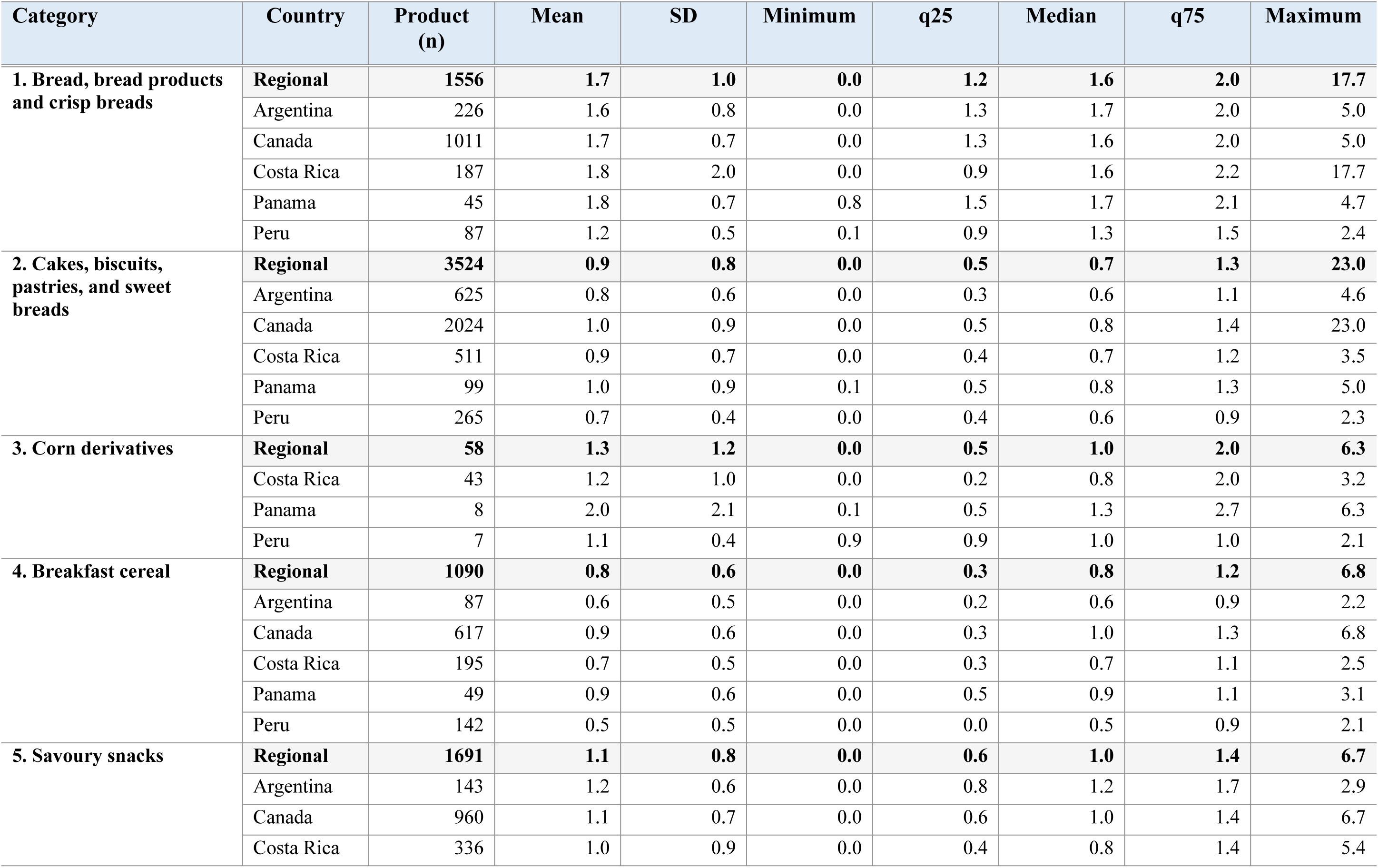
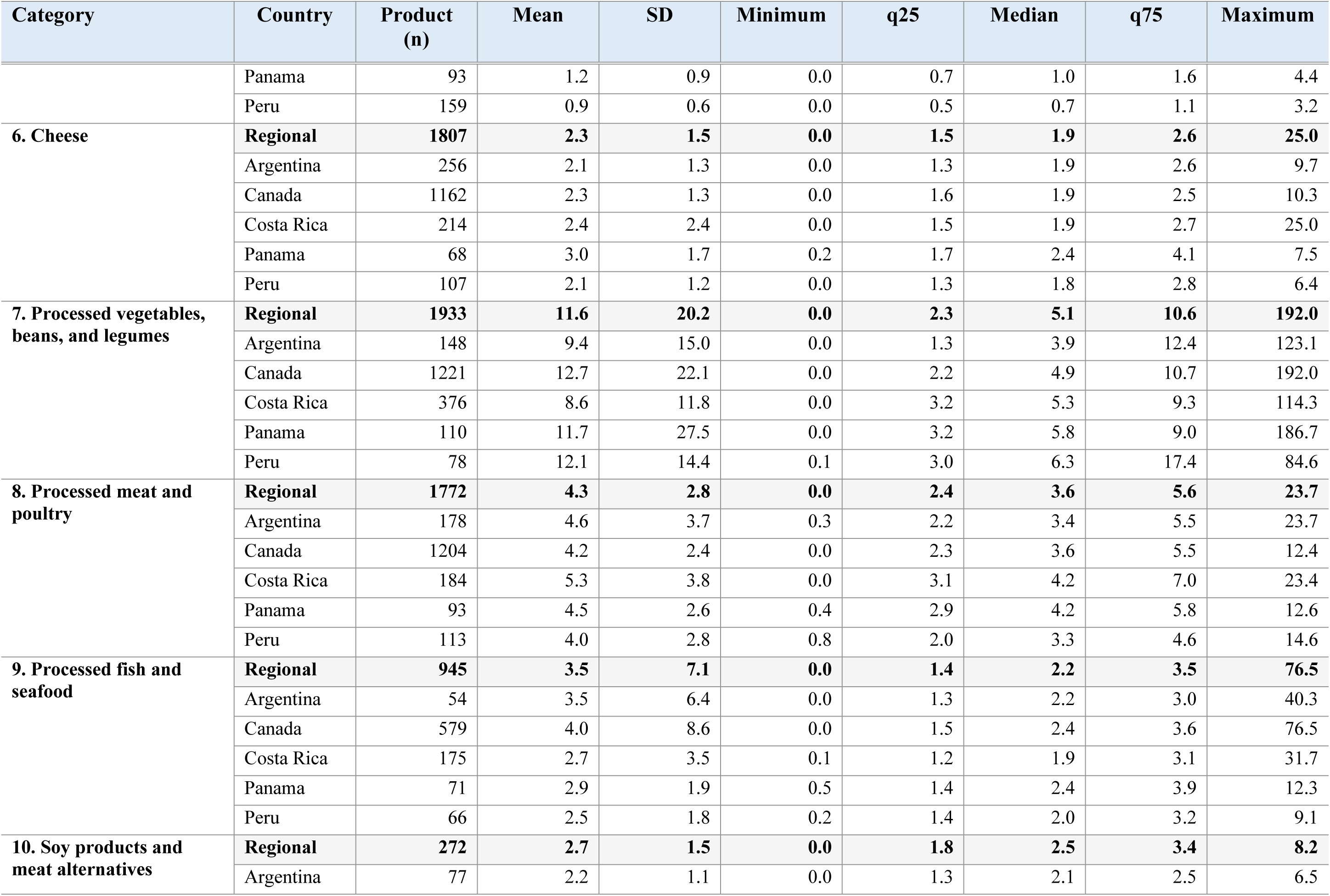
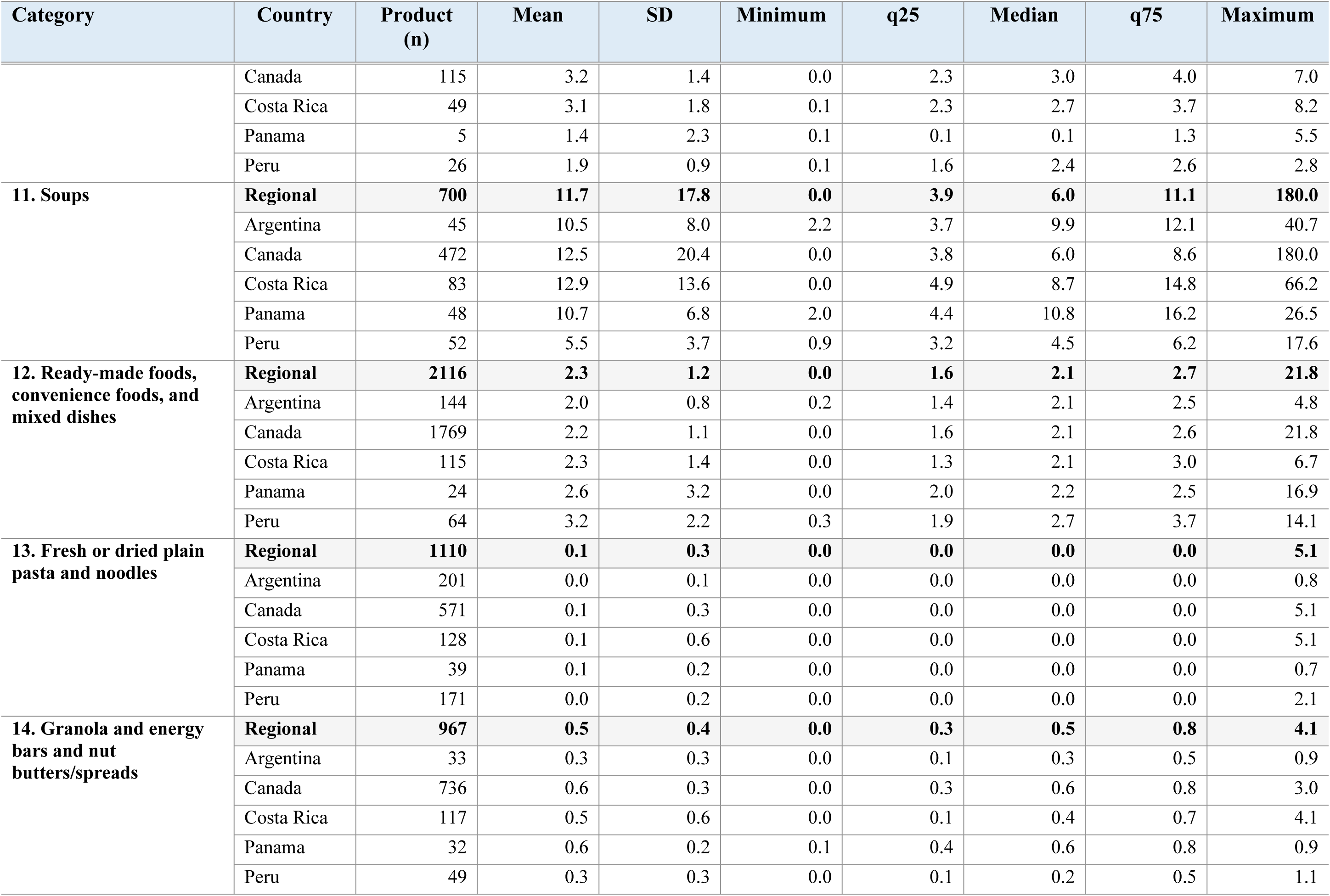
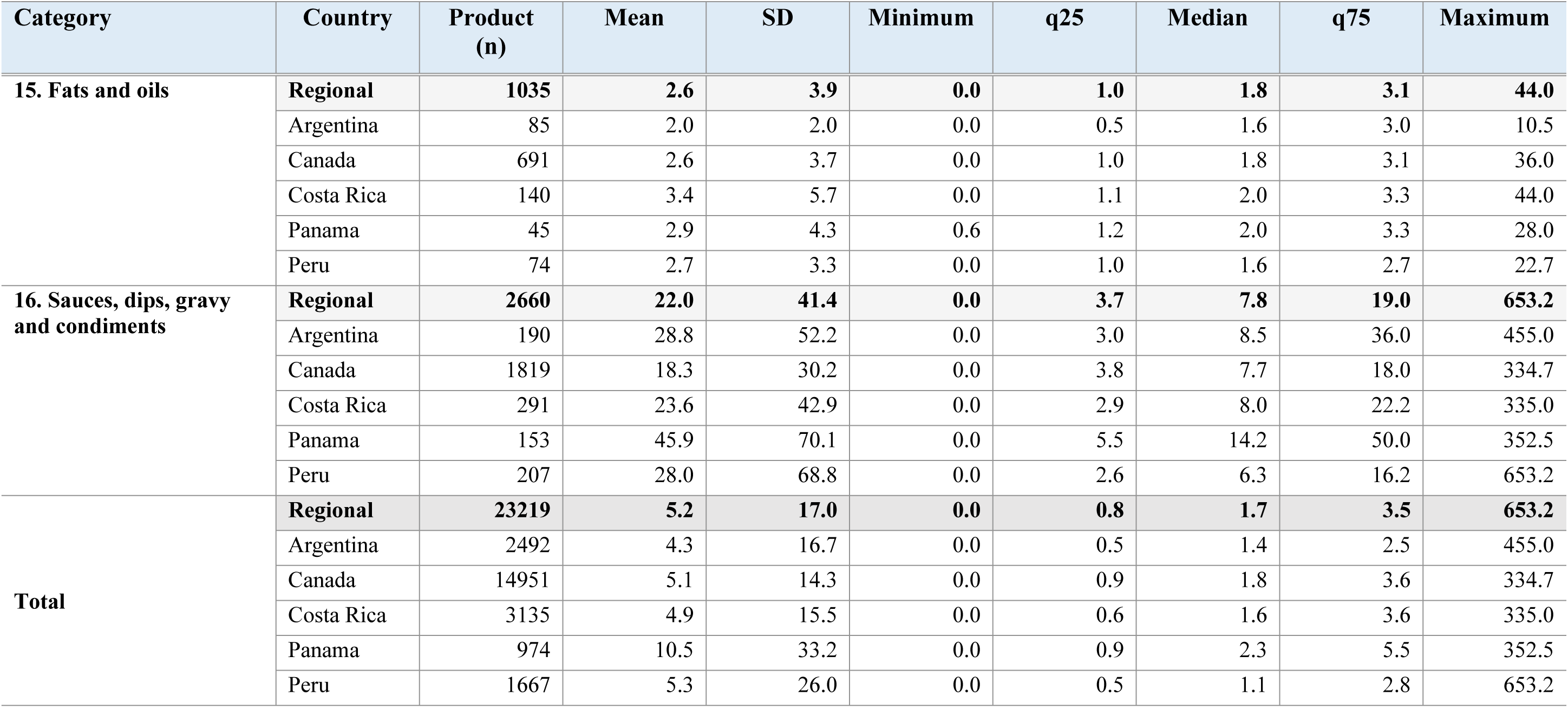
Distribution of sodium content per kcal of packaged foods, by PAHO category at the regional level and by country.

### 3.2 Proportion of foods meeting the 2022 PAHO regional sodium targets by categories and country

Overall, 11,037 of 23,663 (47%) analyzed foods met their respective 2022 PAHO sodium targets (mg/100g). Peru and Argentina had the highest proportion of compliance (52% and 50%, respectively), whereas Panama had the lowest (36%) **(Fig 1).** By food category, the highest proportion of products meeting the regional target was among *‘ready-made foods’* (77%), *‘processed meat and poultry’* (53%) and *‘cheese’* (50%). The lowest proportion of foods meeting the regional target was among *‘bread products’* (30%), *‘granola and energy bars and nut butters/spreads’* (37%) and *‘processed fish and seafood’* (41%). The largest variation between countries was observed in *‘soy products and meat alternatives’* (range: 28% to 85%) and *‘soups’* (range: 31% to 54%) (**Table 3**).

**Fig 1.**
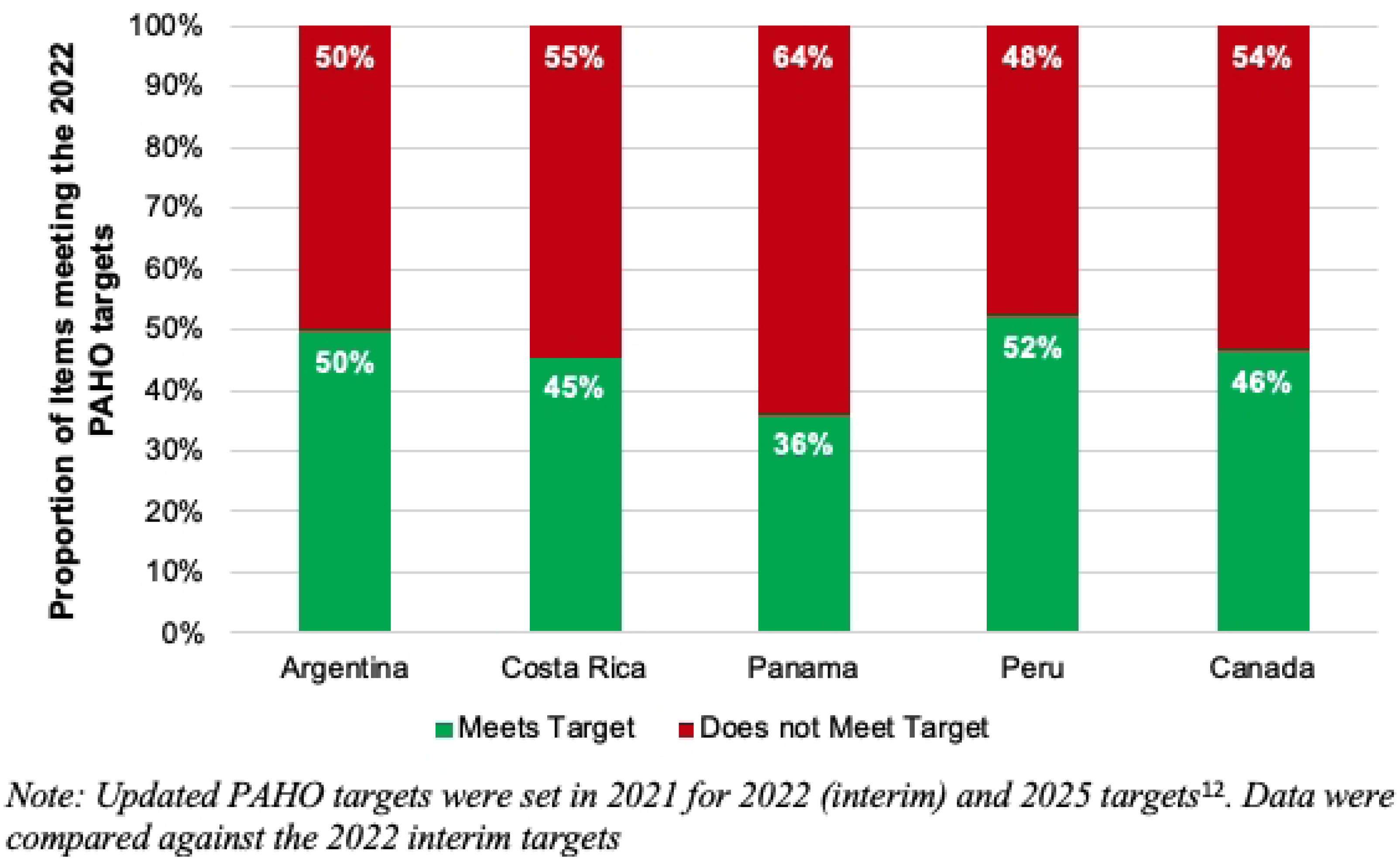
Proportion of products meeting the updated 2022 PAHO Sodium Targets – by country.

**Table 3.**
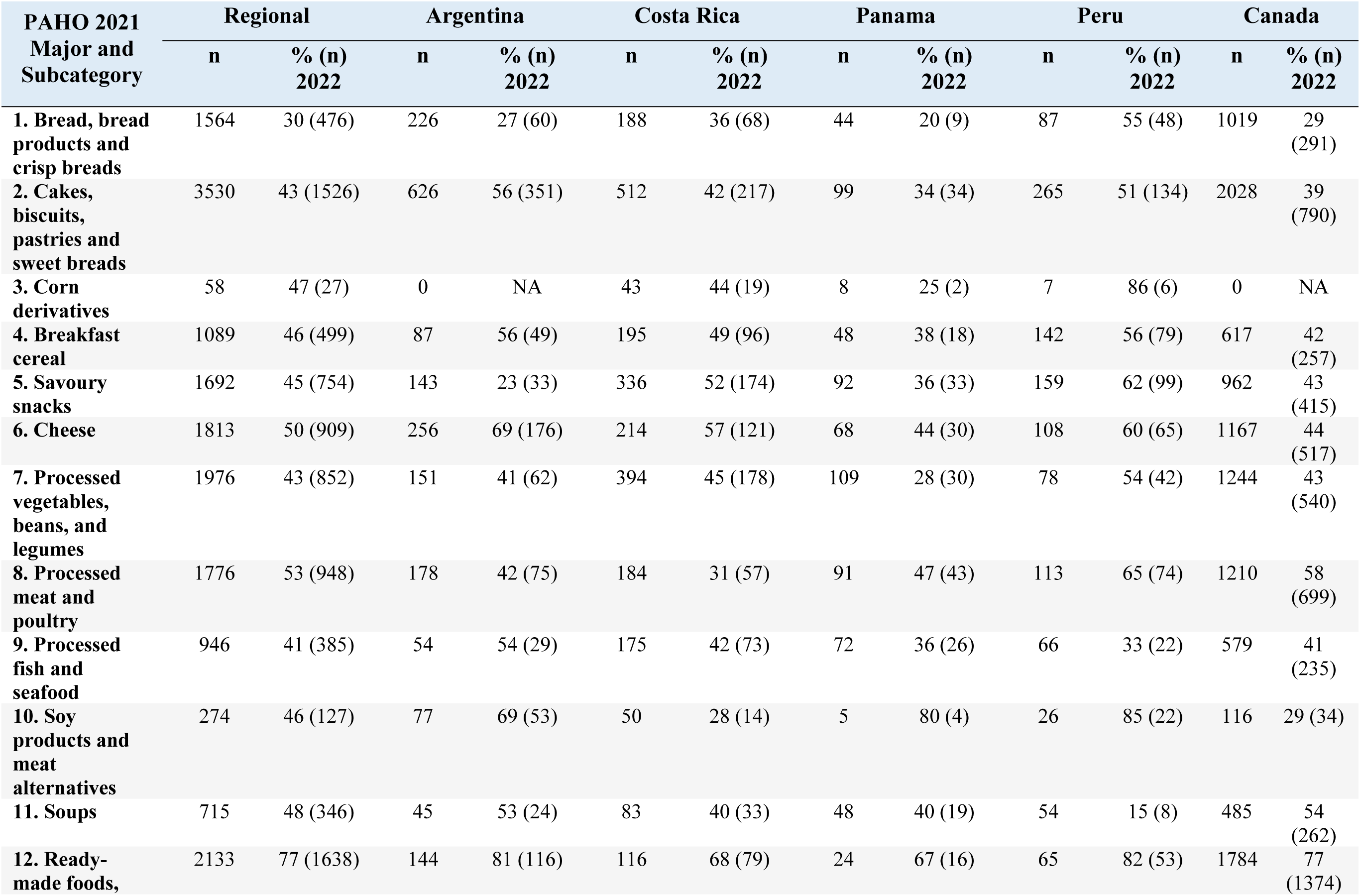
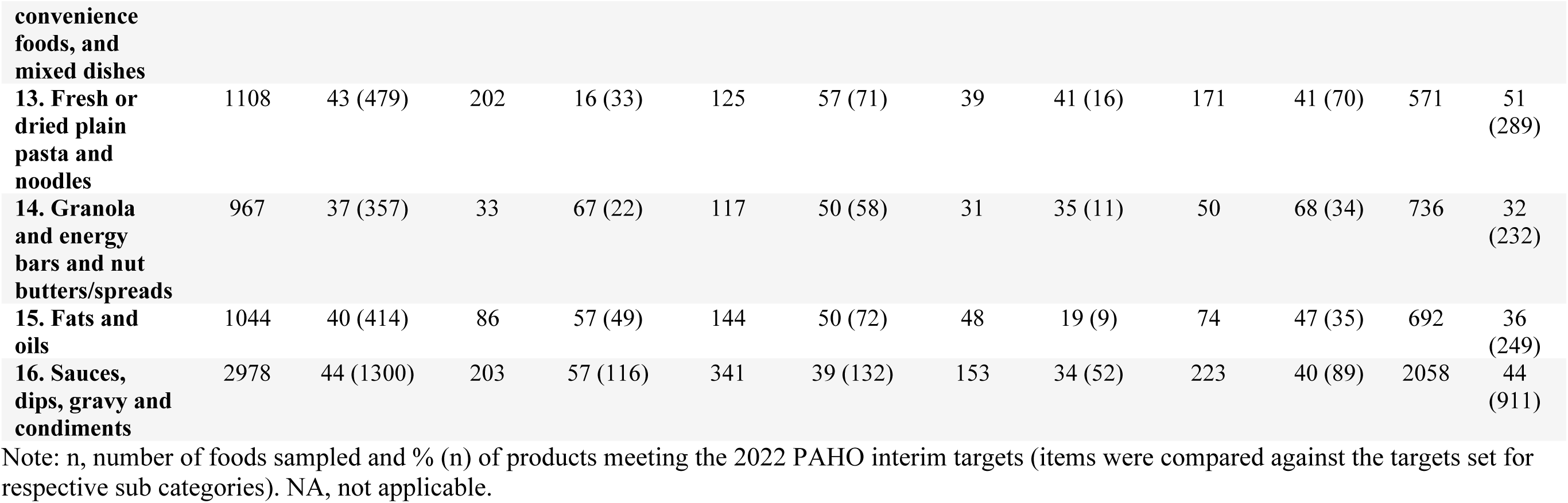
Proportion of products meeting the 2022 PAHO Sodium Interim Targets <u>(mg per 100g/mL)</u>, at the regional and country level, by food category.

With respect to the PAHO targets for sodium levels per kcal, 10,304 of 23,000 (45%) foods met their respective sodium targets (mg/kcal). Peru had the highest proportion of compliance (52%), followed by Costa Rica (48%), whereas Panama only had 37% compliance (**Table 4**). By category, the highest proportion of products meeting the regional target was among *‘savoury snacks*’ (71%), *‘soups’* (67%) and *‘corn derivatives’* (67%). The lowest proportion of foods meeting the regional target was among *‘bread products’* (31%), *‘cheese’* (35%) and *‘ready-made foods’* (35%). The largest variation between countries was observed in ‘soy products and meat alternatives’ (range: 33% to 80%) and ‘fresh or dried plain pasta and noodles’ (range: 16% to 55%) (**Table 4**).

**Table 4.**
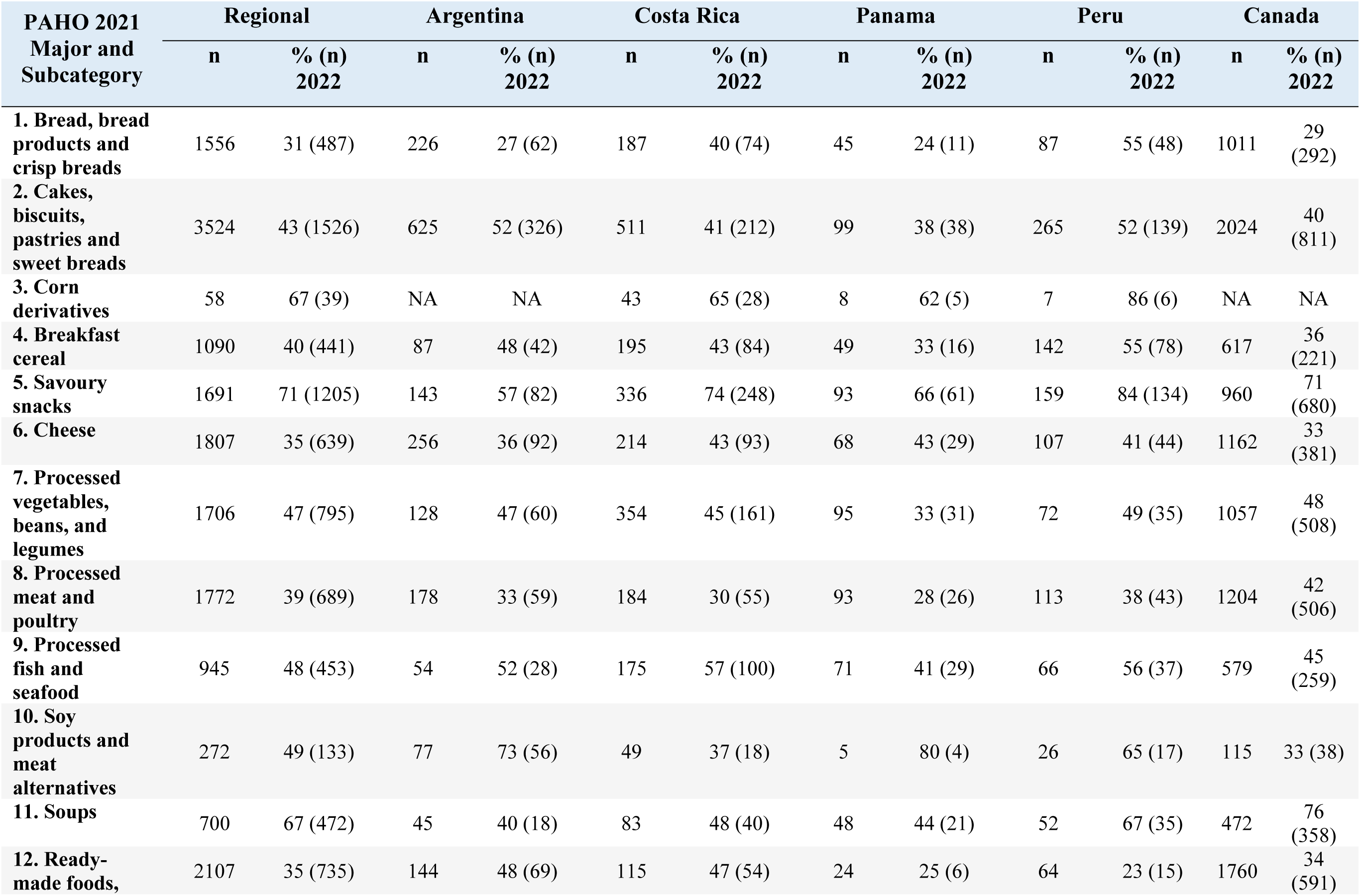
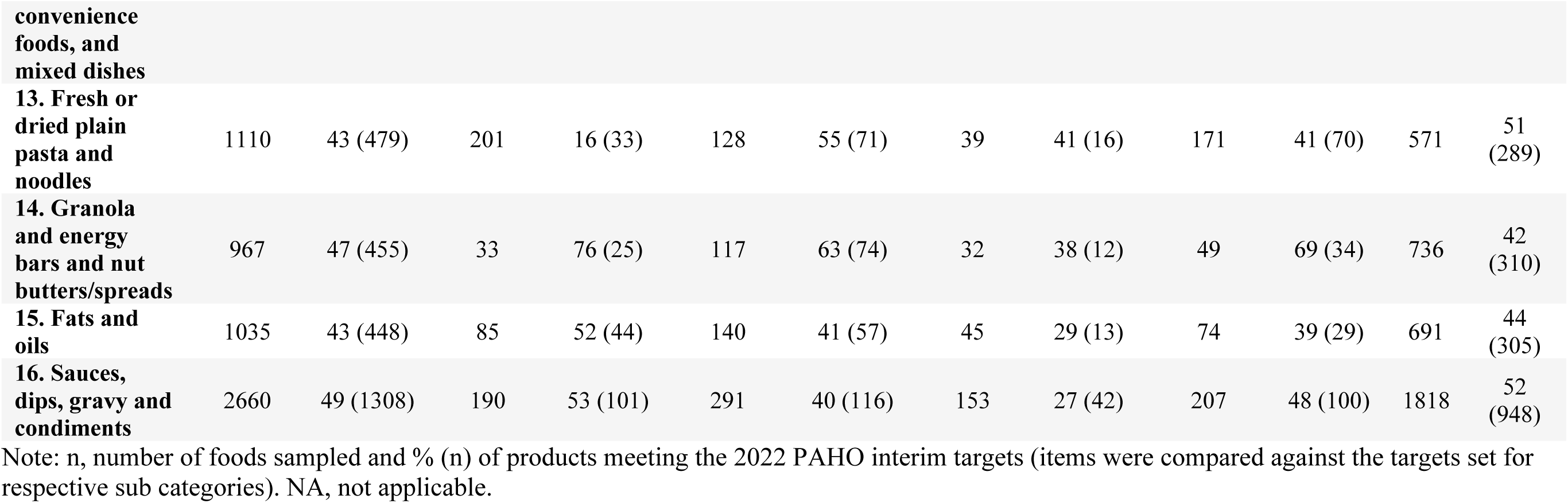
Proportion of products meeting the 2022 PAHO Sodium Targets <u>(mg/kcal)</u>, at the regional and country level, by food category.

**Table 5.** Changes in the proportion of packaged food products meeting the original 2015 PAHO regional sodium reduction targets (2015 PAHO Targets, mg/100g) in PAHO countries between 2015-2016, 2017-2018 and in 2022, by main food category.

| PAHO categories | Year of collection | Products with sodium data (n) | Meeting 2015 Regional Targets (n) | Proportion (%) | Meeting 2015 Lower Targets (n) | Proportion (%) |
| --- | --- | --- | --- | --- | --- | --- |
| **Bread products** | 2015-2016 | 274 | 206 | 75 | 89 | 32 |
|  | 2017-2018 | 218 | 201 | 92 | 99 | 45 |
|  | 2022 | 356 | 296 | 83 | 142 | 40 |
| **Breakfast cereals** | 2015-2016 | 294 | 257 | 87 | 222 | 76 |
|  | 2017-2018 | 299 | 268 | 90 | 237 | 79 |
|  | 2022 | 472 | 452 | 96 | 379 | 80 |
| **Butter and margarine** | 2015-2016 | 84 | 68 | 81 | 43 | 51 |
|  | 2017-2018 | 100 | 82 | 82 | 45 | 45 |
|  | 2022 | 107 | 76 | 71 | 34 | 32 |
| **Cakes** | 2015-2016 | 204 | 142 | 70 | 72 | 35 |
|  | 2017-2018 | 102 | 71 | 70 | 32 | 31 |
|  | 2022 | 358 | 250 | 70 | 96 | 27 |
| **Bouillon cubes and powders** | 2015-2016 | 73 | 38 | 52 | 22 | 30 |
|  | 2017-2018 | 51 | 25 | 49 | 11 | 22 |
|  | 2022 | 46 | 36 | 78 | 34 | 74 |
| **Meat and fish seasonings** | 2015-2016 | 52 | 49 | 94 | 49 | 94 |
|  | 2017-2018 | 64 | 58 | 91 | 58 | 91 |
|  | 2022 | 121 | 105 | 87 | 88 | 73 |
| **Seasonings for side and main dishes** | 2015-2016 | 117 | 117 | 100 | 86 | 74 |
|  | 2017-2018 | 34 | 34 | 100 | 23 | 68 |
|  | 2022 | 43 | 42 | 98 | 8 | 19 |
| **Cookies and sweet cookies** | 2015-2016 | 316 | 294 | 93 | 156 | 49 |
|  | 2017-2018 | 549 | 527 | 96 | 304 | 55 |
|  | 2022 | 857 | 826 | 96 | 499 | 58 |
| **Flavored cookies and crackers** | 2015-2016 | 148 | 141 | 95 | 79 | 53 |
|  | 2017-2018 | 158 | 157 | 99 | 103 | 65 |
|  | 2022 | 273 | 271 | 99 | 163 | 60 |
| **Mayonnaise** | 2015-2016 | 72 | 69 | 96 | 20 | 28 |
|  | 2017-2018 | 80 | 72 | 90 | 16 | 20 |
|  | 2022 | 96 | 89 | 93 | 37 | 39 |
| **Meats and sausages** | 2015-2016 | 269 | 227 | 84 | 67 | 25 |
|  | 2017-2018 | 321 | 260 | 81 | 65 | 20 |
|  | 2022 | 297 | 272 | 92 | 73 | 25 |
| **Cured and preserved meats** | 2015-2016 | 29 | 21 | 72 | 11 | 38 |
|  | 2017-2018 | 62 | 52 | 84 | 24 | 39 |
|  | 2022 | 160 | 131 | 82 | 60 | 38 |
| **Breaded meat and poultry** | 2015-2016 | 64 | 40 | 63 | 10 | 16 |
|  | 2017-2018 | 68 | 56 | 82 | 38 | 56 |
|  | 2022 | 109 | 98 | 90 | 45 | 41 |
| **Pasta and noodles, as consumed** | 2015-2016 | N/A | N/A | N/A | N/A | N/A |
|  | 2017-2018 | 136 | 104 | 76 | 42 | 31 |
|  | 2022 | 111 | 101 | 91 | 69 | 62 |
| **Pasta and noodles, dry uncooked** | 2015-2016 | 217 | 213 | 98 | 206 | 95 |
|  | 2017-2018 | 319 | 318 | 100 | 318 | 100 |
|  | 2022 | 573 | 572 | 100 | 530 | 92 |
| **Snacks** | 2015-2016 | 445 | 344 | 77 | 177 | 40 |
|  | 2017-2018 | 512 | 434 | 85 | 214 | 42 |
|  | 2022 | 730 | 644 | 88 | 358 | 49 |
| **Noodles in broth** | 2015-2016 | 68 | 58 | 85 | 35 | 51 |
|  | 2017-2018 | 42 | 28 | 67 | 15 | 36 |
|  | 2022 | 99 | 64 | 65 | 36 | 36 |
| **Wet and dry soups** | 2015-2016 | 189 | 114 | 60 | 68 | 36 |
|  | 2017-2018 | 126 | 98 | 78 | 71 | 56 |
|  | 2022 | 132 | 113 | 86 | 61 | 46 |
| **Total** | 2015-2016 | 2915 | 2398 | 82 | 1412 | 48 |
|  | 2017-2018 | 3241 | 2845 | 88 | 1715 | 53 |
|  | 2022 | 4940 | 4438 | 90 | 2712 | 55 |

Note: NA, not applicable.

### 3.3 Changes in the proportion of foods meeting the old 2015 PAHO sodium targets

The longitudinal analysis included a total of 10,571 foods, of which 2,915 foods were from the 2015-2016 collection, 3,241 foods were from the 2017-2018 collection, and 4,415 foods were from the 2022 collection (countries: Argentina, Costa Rica, and Peru) across 18 2015 PAHO sodium food categories (**Fig 2**). Overall, the proportion of foods meeting the 2015 PAHO sodium regional target among the three countries was 82, 88 and 90% for 2015-2016, 2017-2018 and 2022, respectively (**Fig 2A**). The proportion of foods meeting the 2015 PAHO lower targets were 48, 53 and 61% for 2015-2016, 2017-2018 and 2022 (**Fig 2B**). There was an increase in the proportion of items meeting the regional targets, with a 9, 9 and 5% increase from 2015-2022 for Argentina, Costa Rica, and Peru, respectively (**Fig 3**).

**Fig 2.**
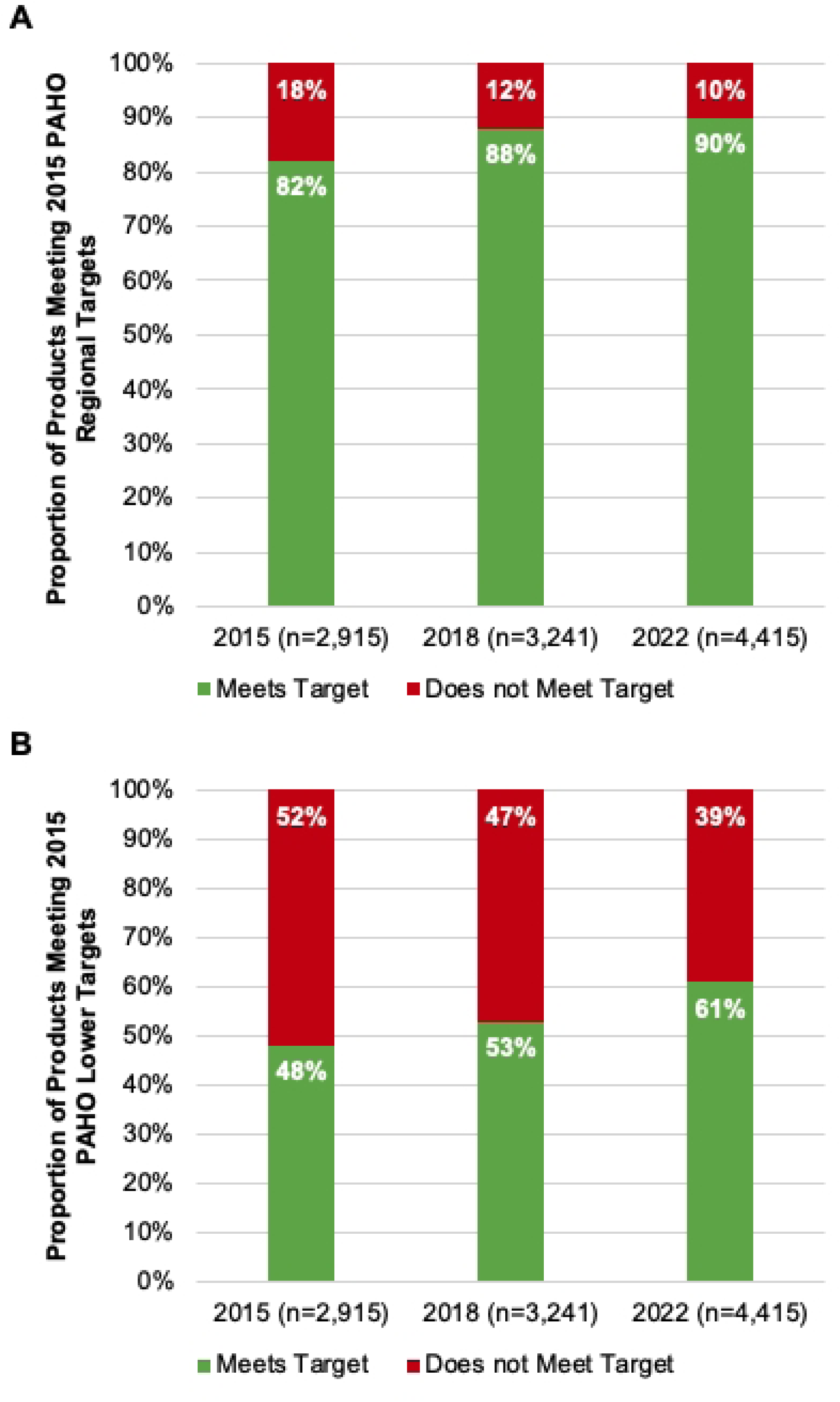
A) Proportion of products meeting the PAHO 2015 Regional Sodium Targets from 2015 to 2022 (Argentina, Costa Rica, Peru) – overall; B) Proportion of products meeting the PAHO 2015 Lower Sodium Targets from 2015 to 2022 (Argentina, Costa Rica, Peru) – overall

**Fig 3.**
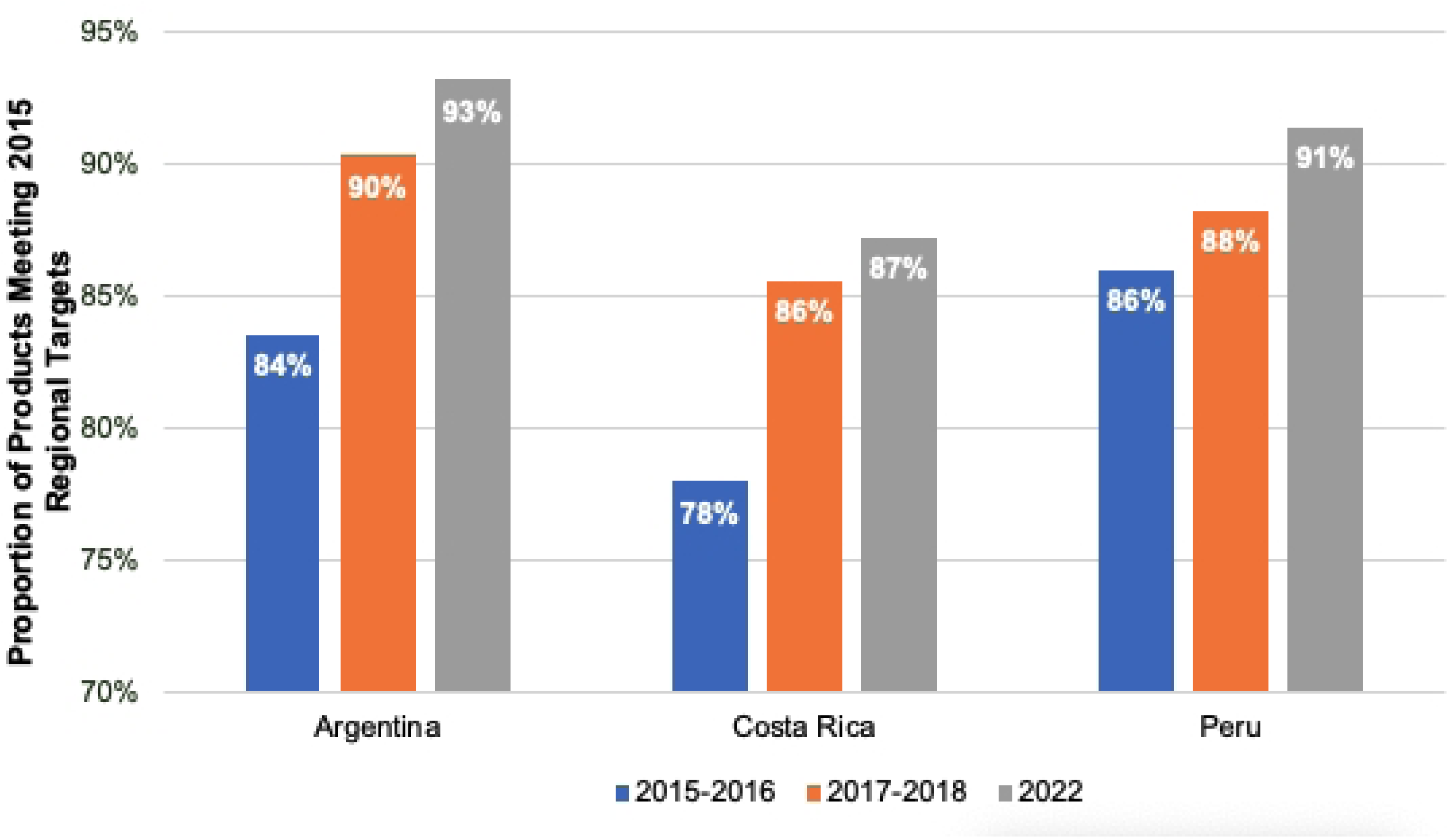
Proportion of products meeting the 2015 PAHO Sodium Targets – by country (2015 – 2022)

## 4. Discussion

This study provides an up-to-date assessment of sodium levels in packaged foods sold in five countries in the Americas, after the publication of the Updated PAHO Regional Sodium Reduction Targets in 2021^12^. The results showed that the proportion of foods meeting the older 2015 PAHO sodium regional targets has gradually increased from 82% to 90%. However, the progress is slow and there still needs to be improvement in compliance with the Updated Sodium Reduction Targets. Further accelerated actions are needed for the achievement of sodium reduction goals. These data shed light into the progress in different countries in the Region towards the current sodium reduction interventions aligned with recommendations from the WHO’s SHAKE technical package for sodium reduction ^15^ and other initiatives to reach the 2025 global target of a 30% relative reduction in mean population intake of sodium.

‘*Processed meat and poultry’* and *‘sauces, dips, gravy, and condiments’* have the highest median sodium levels per 100g. This result is consistent with the previous regional study^9^ that showed a similar median sodium level for *‘processed meats’* and a lower median sodium level in *‘sauces/condiments’*. In addition to the sodium level by weight, sodium density (mg/kcal) accounts for the variation in energy consumption and has been shown to be more strongly associated with blood pressure than total intake^16^. Our results show that *‘sauces, dips, gravy, and condiments’*, *‘soups’* and *‘processed vegetables, beans, and legumes’* had the highest sodium density. These results are expected, as these food categories have lower energy density. Overall, the results also showed large variations within food categories, suggesting that lower sodium options are available among different foods and showing the technical feasibility of reducing sodium levels in these categories. This is important, as it highlights the importance of combining other strategies, such as front-of-pack labels to help consumers identify lower sodium options, social marketing campaigns to encourage consumers to select lower sodium foods, marketing restrictions to products with high sodium contents and nutrition standards for foods and beverages available in schools and other settings, to disincentivize consumption of such products. However, the high average and median sodium levels in many categories also suggested that governments need to encourage sodium reduction at the manufacturer level through food reformulation by setting mandatory sodium reduction targets for key food categories.

When examining the compliance in 2022 with the updated regional targets, almost half of the analyzed foods met their respective sodium targets (mg/100g). This proportion is the same as the previously reported compliance (47%) with the 2015 lower target (which is lower, if not the same, as the 2022 regional target)^9^, suggesting a small increase in the number of foods meeting the targets or the introduction of new low sodium alternative products. The compliance rate between countries was similar, except for Panama (36%). However, this needs careful interpretation due to the different stages at which each country is in its efforts to reduce sodium.

On examination of the longitudinal progress of Argentina, Costa Rica and Peru, there was an 8% increase in compliance when compared to the 2015 regional target (82% to 90%) and an 13% increase when compared to the 2015 lower target (48% to 61%). While this may indicate some progress, it remains insufficient to achieve the target of a 30% relative reduction in mean population intake of sodium by 2025. Therefore, stronger measures and greater compliance with the set targets are needed.

These five countries each have national-level policies supporting sodium reduction. For example, Argentina included sodium maximum levels for certain food groups by passing Act 26905 in 2014 and there were education campaigns and restaurant policies to reduce sodium intake^17^.

There was also ongoing monitoring of Argentina’s sodium content in foods. Studies in 2017-2018 and 2022 showed that over 90% and 94% of surveyed food products, respectively, complied with the national sodium reduction law^18,19^. However, this suggests that the current local limits are too lax compare with the regional targets and require further adjustment; and the necessity to include sodium source food groups that are currently not included, such as cheese and puff pastries^20^.

While Costa Rica has a voluntary sodium reduction strategy to reduce sodium levels in some key food categories, progress has been monitored continuously^21–23^. A study published by Vega-Solano et al. in 2019 showed 87% compliance with the voluntary national sodium targets. In the case of Peru, there is no specific national sodium reduction strategy in place; however, Peru implemented ‘high in’ FOPL regulations in 2019 that requires foods exceeding established thresholds for nutrients of concern (i.e., sugar, saturated fat, trans fat, and sodium) to display a ‘high in’ symbol^24^. This law could indirectly motivate food industry to reformulate products to avoid a ‘high in’ symbol, as already seen in Peru and other countries^25,26^. Canada published a voluntary sodium reduction guideline for prepackaged foods in 2012 and updated the targets in 2020^27,28^. Evaluations of this initiative have shown minimal progress over time, with only 14% of food categories meeting sodium reduction targets in 2017^29^. Panama has recently launched a national plan to reduce sodium intake (2022-2025)^30^.

Overall, these interventions aiming to reduce sodium intake by encouraging food reformulations could have important public health impacts, as several simulation studies found that an important number of CVDs could have been prevented or delayed if the reformulation targets or sodium guidelines would have been met^31–34^. Worth noting that mandatory policies have shown to be more effective than voluntary initiatives^35,36^. For instance, a recent evaluation in South Africa, one of the fewest countries with mandatory sodium levels for key food categories, showed a reduction in salt intake of 1.15 g/day during the first phase of the program (2015-2019), a significant reduction in population-level sodium intake, while other voluntary initiatives in countries like Brazil and Canada have shown only modest results in meeting sodium reduction targets^29,37^. Given the slow progress in meeting regional sodium reduction targets, our results highlight the importance of establishing and monitoring effective and feasible mandatory sodium reduction targets to prevent diet-related NCD deaths.

This study has both strengths and limitations that should be considered when interpreting our findings. Variations in sampling sizes among countries could potentially influence the overall study results. Therefore, a detailed examination of variations within each country at the subcategory level would be necessary, although out of the scope of this analysis. However, we do provide results of progress relative to the PAHO targets, at the major category level for each country. Furthermore, the data in this study were based on Nutrition facts table (NFt) information, with a permitted variation margin of 20% when reporting nutrient values in NFts. Additionally, several products per country were excluded due to the absence of sodium level declarations on food labels, as some countries within the region lack mandatory nutrient values declaration. Regional comparison at different time points was limited to only three countries, including other countries in future analyses will provide a more comprehensive assessment at the regional level. However, this regional analysis benefited from the utilization of common methodologies across countries (e.g., data collection, data cleaning, and analysis). Additionally, several quality assurance measures were performed including food category categorization and validation, outlier check and Atwater validation. Furthermore, prices of products were not included in this study. To ensure the decrease in sodium content is not driven by the lower sodium products that might be higher in price (which likely will not target the majority of population), future studies should also consider including sales-weighted average sodium levels to account for the actual sales of foods sampled.

## 5. Conclusion

Overall, this study provides an ongoing surveillance of the sodium content in package foods sold in five countries in the Americas, under key food categories of the Updated PAHO Regional Sodium Reduction Targets. Around half of the examined foods met their respective sodium targets and there has been some improvement in the compliance overtime, although the reductions have been modest at best. Further efforts are required to reach the WHO’s global sodium reduction goal by 2025, such as implementation of national mandatory sodium reduction targets, reformulation guidelines, FOPL regulations, marketing restrictions, social marketing campaigns, and nutrition standards in schools and other settings, among others. Moreover, there is a need to transform the food systems and promote a healthy diet based on unprocessed or minimally processed foods, rather than ultraprocessed foods.

## Conflict of interest

The authors declare that the research was conducted in the absence of any commercial or financial relationships that could be construed as a potential conflict of interest.

## Authors’ contributions

YY, NF, VT, LG, ABM, HNR, MRA, PAR, MFKL, FDC, MMH, KYS, LSG, LA, LN and MRL conceptualized the study design; YY, NF, VT, LG, ABM, HNR, MRA, PAR, MFKL, FDC, MMH, KYS, LSG, LA, LN and MRL interpreted the findings; YY, NF VT, LG, ABM, HNR, MRA, PAR, MFKL, FDC, MMH, KYS, and LSG conducted the study; YY and NF wrote the original draft; YY performed the statistical analysis. All authors critically reviewed and approved the final manuscript.

## Funding

This research was funded by the Canadian Institutes of Health Research (PJT-152979), Pan American Health Organization (PAHO/WHO) and Resolve to Save Lives.

## Data Availability

All relevant data are within the manuscript and its Supporting Information files. The branded prepackaged food composition database used in this study is from the University of Toronto’s Food Label Information Program for Latin America and Caribbean countries (FLIP-LAC) and FLIP Canada. Any requests to access the FLIP database can be directed to.

## Acknowledgments

The authors would like to express their gratitude to Alyssa Schermel for her support to country teams with the FLIP-LAC application and platform. Also, to country team members Luciana Castronuovo (Argentina) and Ana Atencio (Panama) for their contributions throughout the project.

## Appendix

**Appendix A.**
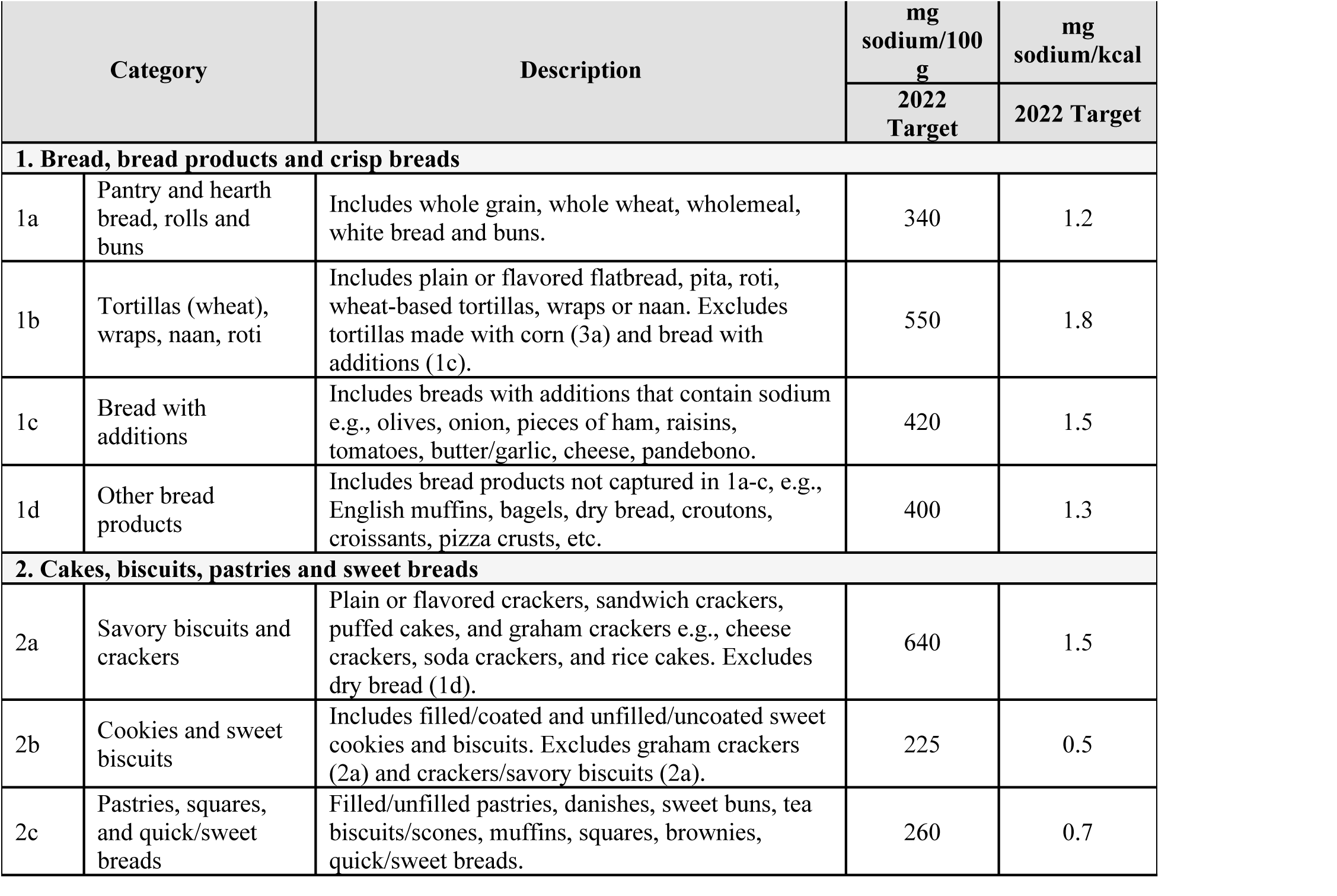
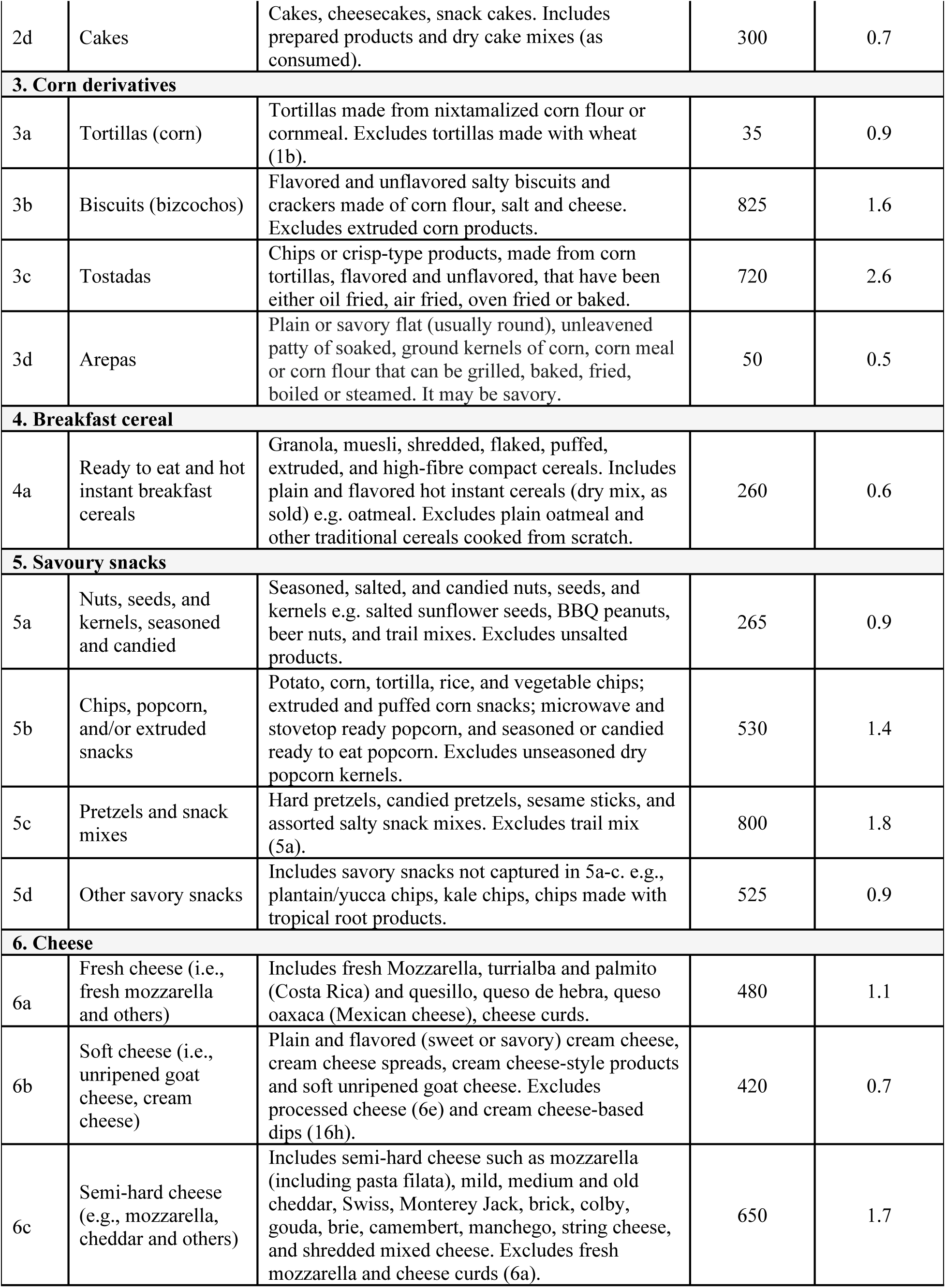
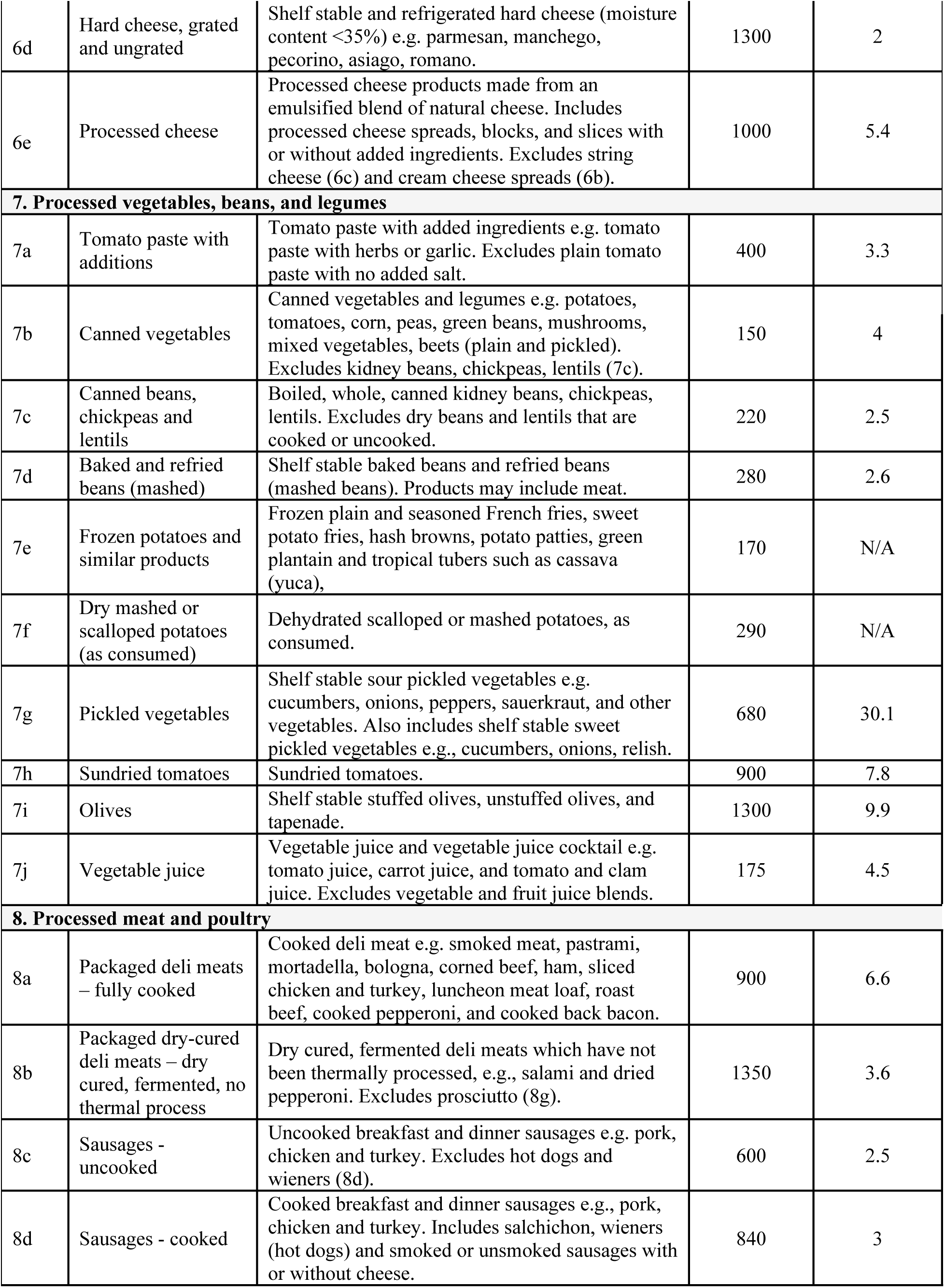
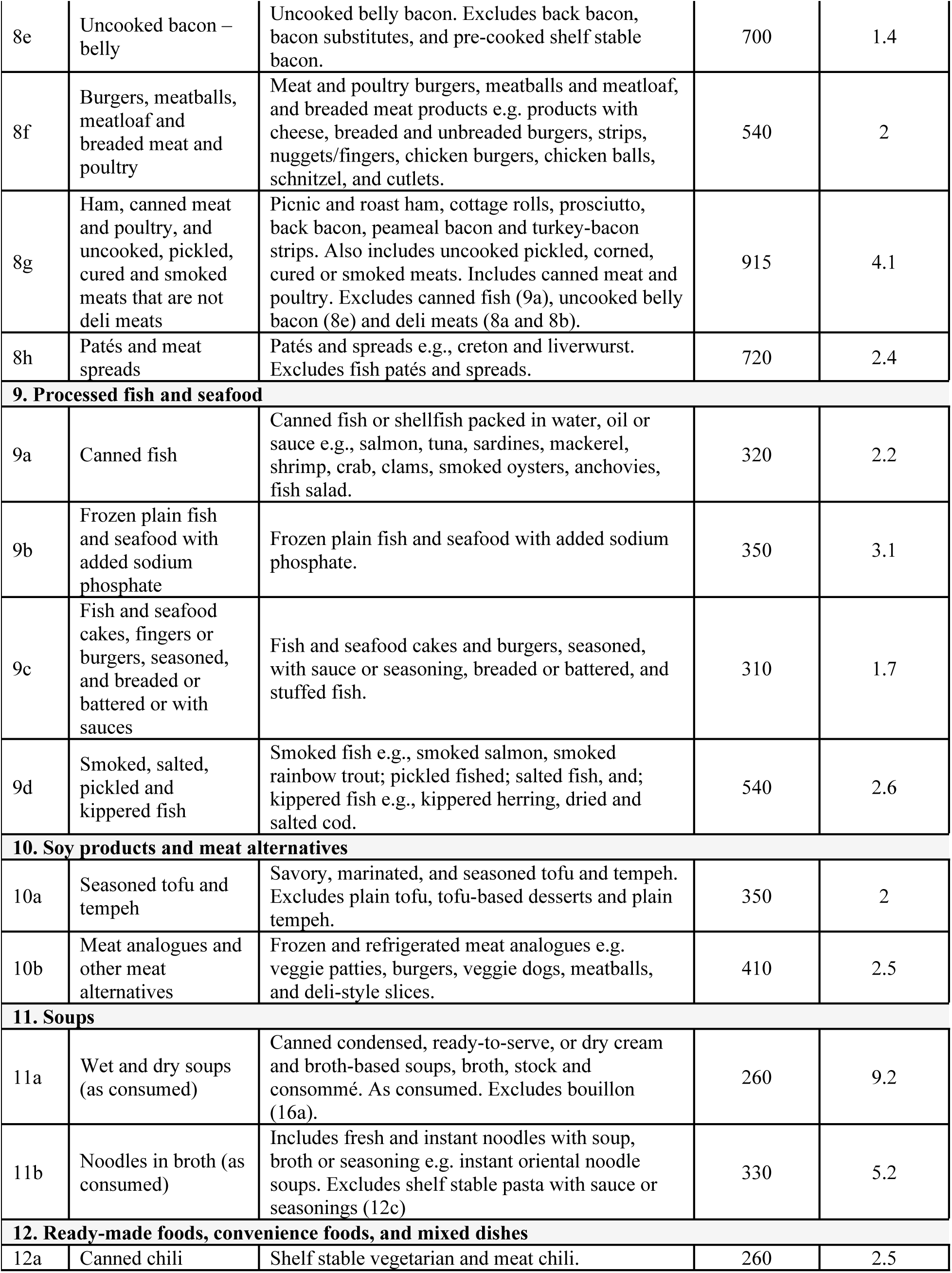
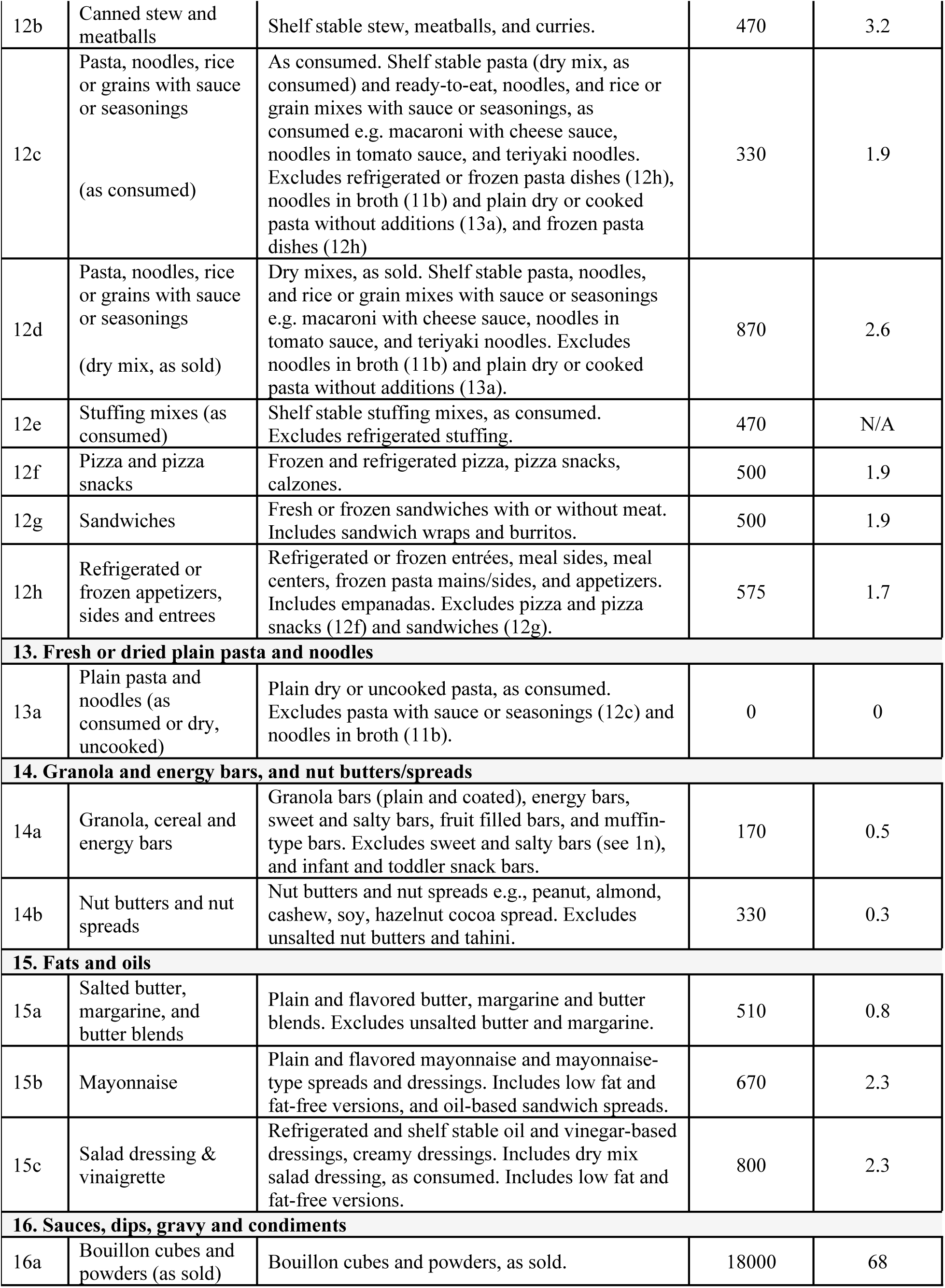
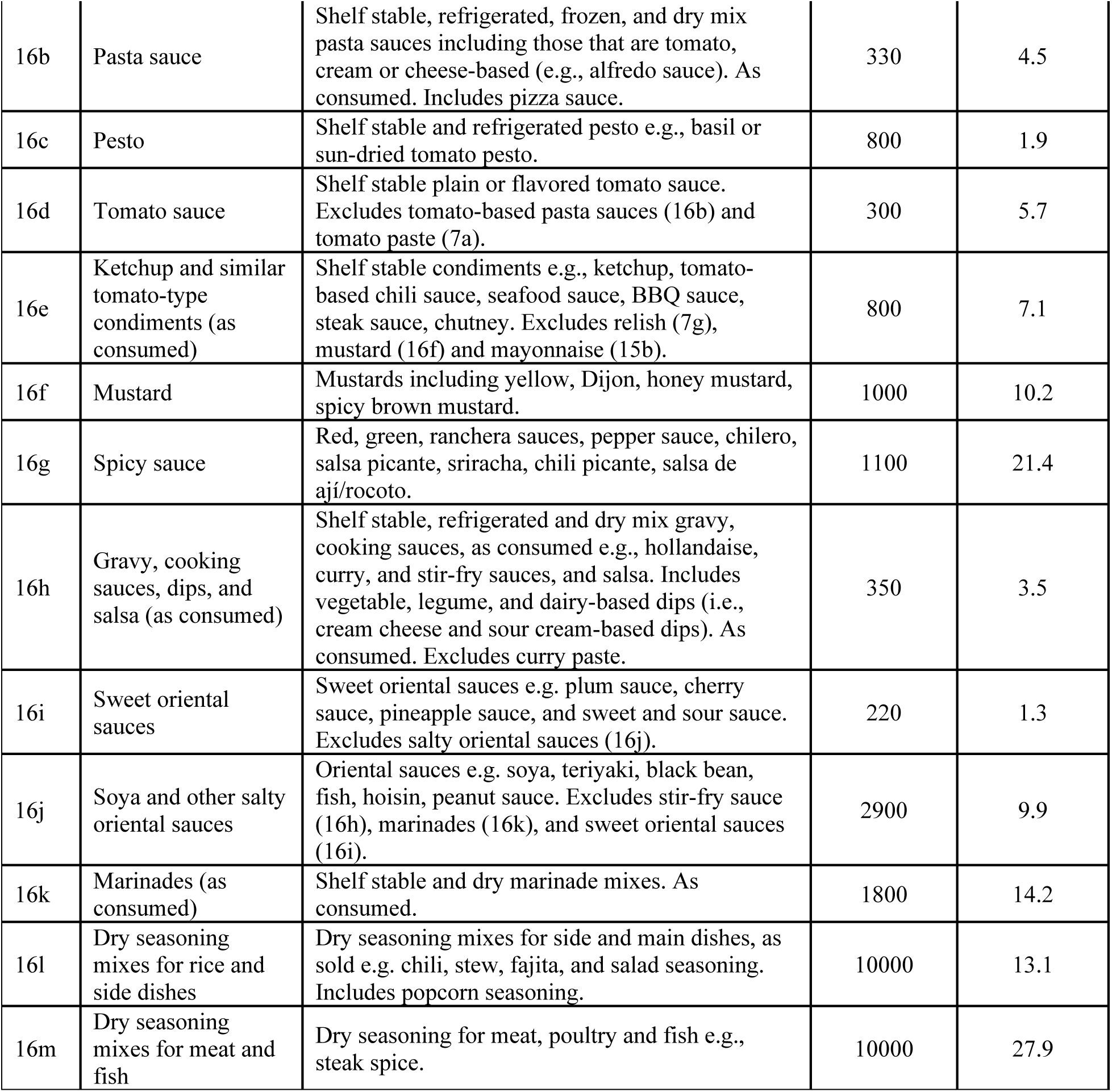
Updated PAHO 2022 Regional Sodium Reduction Targets^12^.

| Category |  | Description | mg sodium/100 g | mg sodium/kcal |
| --- | --- | --- | --- | --- |
|  |  |  | 2022 Target | 2022 Target |
| 1. Bread, bread products and crisp breads |  |  |  |  |
| 1a | Pantry and hearth bread, rolls and buns | Includes whole grain, whole wheat, wholemeal, white bread and buns. | 340 | 1.2 |
| 1b | Tortillas (wheat), wraps, naan, roti | Includes plain or flavored flatbread, pita, roti, wheat-based tortillas, wraps or naan. Excludes tortillas made with corn (3a) and bread with additions (1c). | 550 | 1.8 |
| 1c | Bread with additions | Includes breads with additions that contain sodium e.g., olives, onion, pieces of ham, raisins, tomatoes, butter/garlic, cheese, pandebono. | 420 | 1.5 |
| 1d | Other bread products | Includes bread products not captured in 1a-c, e.g., English muffins, bagels, dry bread, croutons, croissants, pizza crusts, etc. | 400 | 1.3 |
| 2. Cakes, biscuits, pastries and sweet breads |  |  |  |  |
| 2a | Savory biscuits and crackers | Plain or flavored crackers, sandwich crackers, puffed cakes, and graham crackers e.g., cheese crackers, soda crackers, and rice cakes. Excludes dry bread (1d). | 640 | 1.5 |
| 2b | Cookies and sweet biscuits | Includes filled/coated and unfilled/uncoated sweet cookies and biscuits. Excludes graham crackers (2a) and crackers/savory biscuits (2a). | 225 | 0.5 |
| 2c | Pastries, squares, and quick/sweet breads | Filled/unfilled pastries, danishes, sweet buns, tea biscuits/scones, muffins, squares, brownies, quick/sweet breads. | 260 | 0.7 |
| 2d | Cakes | Cakes, cheesecakes, snack cakes. Includes prepared products and dry cake mixes (as consumed). | 300 | 0.7 |
| **3. Corn derivatives** |  |  |  |  |
| 3a | Tortillas (corn) | Tortillas made from nixtamalized corn flour or cornmeal. Excludes tortillas made with wheat (1b). | 35 | 0.9 |
| 3b | Biscuits (bizcochos) | Flavored and unflavored salty biscuits and crackers made of corn flour, salt and cheese. Excludes extruded corn products. | 825 | 1.6 |
| 3c | Tostadas | Chips or crisp-type products, made from corn tortillas, flavored and unflavored, that have been either oil fried, air fried, oven fried or baked. | 720 | 2.6 |
| 3d | Arepas | Plain or savory flat (usually round), unleavened patty of soaked, ground kernels of corn, corn meal or corn flour that can be grilled, baked, fried, boiled or steamed. It may be savory. | 50 | 0.5 |
| **4. Breakfast cereal** |  |  |  |  |
| 4a | Ready to eat and hot instant breakfast cereals | Granola, muesli, shredded, flaked, puffed, extruded, and high-fibre compact cereals. Includes plain and flavored hot instant cereals (dry mix, as sold) e.g. oatmeal. Excludes plain oatmeal and other traditional cereals cooked from scratch. | 260 | 0.6 |
| **5. Savoury snacks** |  |  |  |  |
| 5a | Nuts, seeds, and kernels, seasoned and candied | Seasoned, salted, and candied nuts, seeds, and kernels e.g. salted sunflower seeds, BBQ peanuts, beer nuts, and trail mixes. Excludes unsalted products. | 265 | 0.9 |
| 5b | Chips, popcorn, and/or extruded snacks | Potato, corn, tortilla, rice, and vegetable chips; extruded and puffed corn snacks; microwave and stovetop ready popcorn, and seasoned or candied ready to eat popcorn. Excludes unseasoned dry popcorn kernels. | 530 | 1.4 |
| 5c | Pretzels and snack mixes | Hard pretzels, candied pretzels, sesame sticks, and assorted salty snack mixes. Excludes trail mix (5a). | 800 | 1.8 |
| 5d | Other savory snacks | Includes savory snacks not captured in 5a-c. e.g., plantain/yucca chips, kale chips, chips made with tropical root products. | 525 | 0.9 |
| **6. Cheese** |  |  |  |  |
| 6a | Fresh cheese (i.e., fresh mozzarella and others) | Includes fresh Mozzarella, turrialba and palmito (Costa Rica) and quesillo, queso de hebra, queso oaxaca (Mexican cheese), cheese curds. | 480 | 1.1 |
| 6b | Soft cheese (i.e., unripened goat cheese, cream cheese) | Plain and flavored (sweet or savory) cream cheese, cream cheese spreads, cream cheese-style products and soft unripened goat cheese. Excludes processed cheese (6e) and cream cheese-based dips (16h). | 420 | 0.7 |
| 6c | Semi-hard cheese (e.g., mozzarella, cheddar and others) | Includes semi-hard cheese such as mozzarella (including pasta filata), mild, medium and old cheddar, Swiss, Monterey Jack, brick, colby, gouda, brie, camembert, manchego, string cheese, and shredded mixed cheese. Excludes fresh mozzarella and cheese curds (6a). | 650 | 1.7 |
| 6d | Hard cheese, grated and ungrated | Shelf stable and refrigerated hard cheese (moisture content <35%) e.g. parmesan, manchego, pecorino, asiago, romano. | 1300 | 2 |
| 6e | Processed cheese | Processed cheese products made from an emulsified blend of natural cheese. Includes processed cheese spreads, blocks, and slices with or without added ingredients. Excludes string cheese (6c) and cream cheese spreads (6b). | 1000 | 5.4 |
| **7. Processed vegetables, beans, and legumes** |  |  |  |  |
| 7a | Tomato paste with additions | Tomato paste with added ingredients e.g. tomato paste with herbs or garlic. Excludes plain tomato paste with no added salt. | 400 | 3.3 |
| 7b | Canned vegetables | Canned vegetables and legumes e.g. potatoes, tomatoes, corn, peas, green beans, mushrooms, mixed vegetables, beets (plain and pickled). Excludes kidney beans, chickpeas, lentils (7c). | 150 | 4 |
| 7c | Canned beans, chickpeas and lentils | Boiled, whole, canned kidney beans, chickpeas, lentils. Excludes dry beans and lentils that are cooked or uncooked. | 220 | 2.5 |
| 7d | Baked and refried beans (mashed) | Shelf stable baked beans and refried beans (mashed beans). Products may include meat. | 280 | 2.6 |
| 7e | Frozen potatoes and similar products | Frozen plain and seasoned French fries, sweet potato fries, hash browns, potato patties, green plantain and tropical tubers such as cassava (yuca), | 170 | N/A |
| 7f | Dry mashed or scalloped potatoes (as consumed) | Dehydrated scalloped or mashed potatoes, as consumed. | 290 | N/A |
| 7g | Pickled vegetables | Shelf stable sour pickled vegetables e.g. cucumbers, onions, peppers, sauerkraut, and other vegetables. Also includes shelf stable sweet pickled vegetables e.g., cucumbers, onions, relish. | 680 | 30.1 |
| 7h | Sundried tomatoes | Sundried tomatoes. | 900 | 7.8 |
| 7i | Olives | Shelf stable stuffed olives, unstuffed olives, and tapenade. | 1300 | 9.9 |
| 7j | Vegetable juice | Vegetable juice and vegetable juice cocktail e.g. tomato juice, carrot juice, and tomato and clam juice. Excludes vegetable and fruit juice blends. | 175 | 4.5 |
| **8. Processed meat and poultry** |  |  |  |  |
| 8a | Packaged deli meats – fully cooked | Cooked deli meat e.g. smoked meat, pastrami, mortadella, bologna, corned beef, ham, sliced chicken and turkey, luncheon meat loaf, roast beef, cooked pepperoni, and cooked back bacon. | 900 | 6.6 |
| 8b | Packaged dry-cured deli meats – dry cured, fermented, no thermal process | Dry cured, fermented deli meats which have not been thermally processed, e.g., salami and dried pepperoni. Excludes prosciutto (8g). | 1350 | 3.6 |
| 8c | Sausages - uncooked | Uncooked breakfast and dinner sausages e.g. pork, chicken and turkey. Excludes hot dogs and wieners (8d). | 600 | 2.5 |
| 8d | Sausages - cooked | Cooked breakfast and dinner sausages e.g., pork, chicken and turkey. Includes salchichon, wieners (hot dogs) and smoked or unsmoked sausages with or without cheese. | 840 | 3 |
| 8e | Uncooked bacon – belly | Uncooked belly bacon. Excludes back bacon, bacon substitutes, and pre-cooked shelf stable bacon. | 700 | 1.4 |
| 8f | Burgers, meatballs, meatloaf and breaded meat and poultry | Meat and poultry burgers, meatballs and meatloaf, and breaded meat products e.g. products with cheese, breaded and unbreaded burgers, strips, nuggets/fingers, chicken burgers, chicken balls, schnitzel, and cutlets. | 540 | 2 |
| 8g | Ham, canned meat and poultry, and uncooked, pickled, cured and smoked meats that are not deli meats | Picnic and roast ham, cottage rolls, prosciutto, back bacon, peameal bacon and turkey-bacon strips. Also includes uncooked pickled, corned, cured or smoked meats. Includes canned meat and poultry. Excludes canned fish (9a), uncooked belly bacon (8e) and deli meats (8a and 8b). | 915 | 4.1 |
| 8h | Patés and meat spreads | Patés and spreads e.g., creton and liverwurst. Excludes fish patés and spreads. | 720 | 2.4 |
| **9. Processed fish and seafood** |  |  |  |  |
| 9a | Canned fish | Canned fish or shellfish packed in water, oil or sauce e.g., salmon, tuna, sardines, mackerel, shrimp, crab, clams, smoked oysters, anchovies, fish salad. | 320 | 2.2 |
| 9b | Frozen plain fish and seafood with added sodium phosphate | Frozen plain fish and seafood with added sodium phosphate. | 350 | 3.1 |
| 9c | Fish and seafood cakes, fingers or burgers, seasoned, and breaded or battered or with sauces | Fish and seafood cakes and burgers, seasoned, with sauce or seasoning, breaded or battered, and stuffed fish. | 310 | 1.7 |
| 9d | Smoked, salted, pickled and kippered fish | Smoked fish e.g., smoked salmon, smoked rainbow trout; pickled fish; salted fish, and; kippered fish e.g., kippered herring, dried and salted cod. | 540 | 2.6 |
| **10. Soy products and meat alternatives** |  |  |  |  |
| 10a | Seasoned tofu and tempeh | Savory, marinated, and seasoned tofu and tempeh. Excludes plain tofu, tofu-based desserts and plain tempeh. | 350 | 2 |
| 10b | Meat analogues and other meat alternatives | Frozen and refrigerated meat analogues e.g. veggie patties, burgers, veggie dogs, meatballs, and deli-style slices. | 410 | 2.5 |
| **11. Soups** |  |  |  |  |
| 11a | Wet and dry soups (as consumed) | Canned condensed, ready-to-serve, or dry cream and broth-based soups, broth, stock and consommé. As consumed. Excludes bouillon (16a). | 260 | 9.2 |
| 11b | Noodles in broth (as consumed) | Includes fresh and instant noodles with soup, broth or seasoning e.g. instant oriental noodle soups. Excludes shelf stable pasta with sauce or seasonings (12c) | 330 | 5.2 |
| **12. Ready-made foods, convenience foods, and mixed dishes** |  |  |  |  |
| 12a | Canned chili | Shelf stable vegetarian and meat chili. | 260 | 2.5 |
| 12b | Canned stew and meatballs | Shelf stable stew, meatballs, and curries. | 470 | 3.2 |
| 12c | Pasta, noodles, rice or grains with sauce or seasonings<br>(as consumed) | As consumed. Shelf stable pasta (dry mix, as consumed) and ready-to-eat, noodles, and rice or grain mixes with sauce or seasonings, as consumed e.g. macaroni with cheese sauce, noodles in tomato sauce, and teriyaki noodles. Excludes refrigerated or frozen pasta dishes (12h), noodles in broth (11b) and plain dry or cooked pasta without additions (13a), and frozen pasta dishes (12h) | 330 | 1.9 |
| 12d | Pasta, noodles, rice or grains with sauce or seasonings<br>(dry mix, as sold) | Dry mixes, as sold. Shelf stable pasta, noodles, and rice or grain mixes with sauce or seasonings e.g. macaroni with cheese sauce, noodles in tomato sauce, and teriyaki noodles. Excludes noodles in broth (11b) and plain dry or cooked pasta without additions (13a). | 870 | 2.6 |
| 12e | Stuffing mixes (as consumed) | Shelf stable stuffing mixes, as consumed. Excludes refrigerated stuffing. | 470 | N/A |
| 12f | Pizza and pizza snacks | Frozen and refrigerated pizza, pizza snacks, calzones. | 500 | 1.9 |
| 12g | Sandwiches | Fresh or frozen sandwiches with or without meat. Includes sandwich wraps and burritos. | 500 | 1.9 |
| 12h | Refrigerated or frozen appetizers, sides and entrees | Refrigerated or frozen entrées, meal sides, meal centers, frozen pasta mains/sides, and appetizers. Includes empanadas. Excludes pizza and pizza snacks (12f) and sandwiches (12g). | 575 | 1.7 |
| **13. Fresh or dried plain pasta and noodles** |  |  |  |  |
| 13a | Plain pasta and noodles (as consumed or dry, uncooked) | Plain dry or uncooked pasta, as consumed. Excludes pasta with sauce or seasonings (12c) and noodles in broth (11b). | 0 | 0 |
| **14. Granola and energy bars, and nut butters/spreads** |  |  |  |  |
| 14a | Granola, cereal and energy bars | Granola bars (plain and coated), energy bars, sweet and salty bars, fruit filled bars, and muffin-type bars. Excludes sweet and salty bars (see 1n), and infant and toddler snack bars. | 170 | 0.5 |
| 14b | Nut butters and nut spreads | Nut butters and nut spreads e.g., peanut, almond, cashew, soy, hazelnut cocoa spread. Excludes unsalted nut butters and tahini. | 330 | 0.3 |
| **15. Fats and oils** |  |  |  |  |
| 15a | Salted butter, margarine, and butter blends | Plain and flavored butter, margarine and butter blends. Excludes unsalted butter and margarine. | 510 | 0.8 |
| 15b | Mayonnaise | Plain and flavored mayonnaise and mayonnaise-type spreads and dressings. Includes low fat and fat-free versions, and oil-based sandwich spreads. | 670 | 2.3 |
| 15c | Salad dressing & vinaigrette | Refrigerated and shelf stable oil and vinegar-based dressings, creamy dressings. Includes dry mix salad dressing, as consumed. Includes low fat and fat-free versions. | 800 | 2.3 |
| **16. Sauces, dips, gravy and condiments** |  |  |  |  |
| 16a | Bouillon cubes and powders (as sold) | Bouillon cubes and powders, as sold. | 18000 | 68 |
| 16b | Pasta sauce | Shelf stable, refrigerated, frozen, and dry mix pasta sauces including those that are tomato, cream or cheese-based (e.g., alfredo sauce). As consumed. Includes pizza sauce. | 330 | 4.5 |
| 16c | Pesto | Shelf stable and refrigerated pesto e.g., basil or sun-dried tomato pesto. | 800 | 1.9 |
| 16d | Tomato sauce | Shelf stable plain or flavored tomato sauce. Excludes tomato-based pasta sauces (16b) and tomato paste (7a). | 300 | 5.7 |
| 16e | Ketchup and similar tomato-type condiments (as consumed) | Shelf stable condiments e.g., ketchup, tomato-based chili sauce, seafood sauce, BBQ sauce, steak sauce, chutney. Excludes relish (7g), mustard (16f) and mayonnaise (15b). | 800 | 7.1 |
| 16f | Mustard | Mustards including yellow, Dijon, honey mustard, spicy brown mustard. | 1000 | 10.2 |
| 16g | Spicy sauce | Red, green, ranchera sauces, pepper sauce, chilero, salsa picante, sriracha, chili picante, salsa de aji/rocoto. | 1100 | 21.4 |
| 16h | Gravy, cooking sauces, dips, and salsa (as consumed) | Shelf stable, refrigerated and dry mix gravy, cooking sauces, as consumed e.g., hollandaise, curry, and stir-fry sauces, and salsa. Includes vegetable, legume, and dairy-based dips (i.e., cream cheese and sour cream-based dips). As consumed. Excludes curry paste. | 350 | 3.5 |
| 16i | Sweet oriental sauces | Sweet oriental sauces e.g. plum sauce, cherry sauce, pineapple sauce, and sweet and sour sauce. Excludes salty oriental sauces (16j). | 220 | 1.3 |
| 16j | Soya and other salty oriental sauces | Oriental sauces e.g. soya, teriyaki, black bean, fish, hoisin, peanut sauce. Excludes stir-fry sauce (16h), marinades (16k), and sweet oriental sauces (16i). | 2900 | 9.9 |
| 16k | Marinades (as consumed) | Shelf stable and dry marinade mixes. As consumed. | 1800 | 14.2 |
| 16l | Dry seasoning mixes for rice and side dishes | Dry seasoning mixes for side and main dishes, as sold e.g. chili, stew, fajita, and salad seasoning. Includes popcorn seasoning. | 10000 | 13.1 |
| 16m | Dry seasoning mixes for meat and fish | Dry seasoning for meat, poultry and fish e.g., steak spice. | 10000 | 27.9 |

*Note: Updated PAHO targets were set in 2021 for 2022 (interim) and 2025 targets<sup>12</sup>. Data were compared against the 2022 interim targets*

## Notes

### Competing Interest Statement

The authors have declared no competing interest.

### Funding Statement

Yes

